# BaySyn: Bayesian Evidence Synthesis for Multi-system Multiomic Integration

**DOI:** 10.1101/2022.08.16.22278812

**Authors:** Rupam Bhattacharyya, Nicholas Henderson, Veerabhadran Baladandayuthapani

**Affiliations:** Department of Biostatistics, University of Michigan, Ann Arbor Michigan 48109, USA

**Keywords:** Additive Gaussian processes, cancer driver genes, gene-drug associations, hierarchical Bayesian variable selection, KEGG gene sets, spike-and-slab priors

## Abstract

The discovery of cancer drivers and drug targets are often limited to the biological systems - from cancer model systems to patients. While multiomic patient databases have sparse drug response data, cancer model systems databases, despite covering a broad range of pharmacogenomic platforms, provide lower lineage-specific sample sizes, resulting in reduced statistical power to detect both functional driver genes and their associations with drug sensitivity profiles. Hence, integrating evidence across model systems, taking into account the pros and cons of each system, in addition to multiomic integration, can more efficiently deconvolve cellular mechanisms of cancer as well as learn therapeutic associations. To this end, we propose *BaySyn* - a hierarchical <u>Bay</u>esian evidence <u>syn</u>thesis framework for multi-system multiomic integration. BaySyn detects functionally relevant driver genes based on their associations with upstream regulators using additive Gaussian process models and uses this evidence to calibrate Bayesian variable selection models in the (drug) outcome layer. We apply BaySyn to multiomic cancer cell line and patient datasets from the Cancer Cell Line Encyclopedia and The Cancer Genome Atlas, respectively, across pan-gynecological cancers. Our mechanistic models implicate several relevant functional genes across cancers such as PTPN6 and ERBB2 in the KEGG adherens junction gene set. Furthermore, our outcome model is able to make higher number of discoveries in drug response models than its uncalibrated counterparts under the same thresholds of Type I error control, including detection of known lineage-specific biomarker associations such as BCL11A in breast and FGFRL1 in ovarian cancers. All our results and implementation codes are freely available via an interactive R Shiny dashboard.

## 1. Introduction

With the advent of sophisticated techniques and platforms, large-scale datasets covering multiple layers of cellular omics are becoming increasingly available.^1,2^ Consistent advancements have been made in the last few years towards adding more dimensions to these high-throughput datasets, namely (1) additional to patient-level disease databases, model systems such as cell lines, patient-derived xenografts and organoids are being studied extensively in context of cancer and other diseases;^3,4^ (2) assessing clinical information and therapeutic response with omics data to make pharmacogenomic discoveries is becoming increasingly common.^5,6^ Multiple challenges arise during investigations of such datasets, including but not limited to computational inefficiency, complex nature of associations among the omic variables considered, and the biological interpretability and clinical implications of the results.^7^ Specifically in context of cancer, the necessity to not only detect biomarker associations with drug/treatment regimens but also to assess the functional relevance and mechanism of such associations is paramount, potentially guiding future therapeutic advances. Thus, novel algorithms that integrate multi-omics patient and model systems profiles can potentially reveal novel biomarkers, drug targets and predictive models in cancer.

### Multi-dimensional data integration in cancer

To address the wide range of complexity and variability in both detection and management of cancer, a number of multi-omics approaches have been able to uncover intricate molecular mechanisms and discover prognostic candidates.^8^ Data integration approaches have proven particularly useful - both vertical (multiple experiments on a common cohort of samples) and horizontal (meta-analysis of different cohorts) integration methods have been developed.^9^ To simultaneously identify pharmacogenomic associations and corresponding functional mechanisms, singular usage of either of these dimensions is insufficient due to the richness of the currently available omics databases. Multi-omics patient databases of cancer such as The Cancer Genome Atlas (TCGA),^10^ while rich in transcriptomic, proteomic and other levels of omics profiles, do not typically provide comprehensive and systematic drug response on the same cohort of patients, restricting utilization of these profiles directly in pharmacogenomic contexts. Model systems databases such as the Cancer Cell Line Encyclopedia (CCLE)^11^ and Genomics of Drug Sensitivity in Cancer (GDSC)^12^ provide both molecular profiles and drug sensitivity information on the same set of models, but the cancer- or lineage-specific sample sizes of such databases are lower than their patient counterparts and association models built solely on them may suffer from the lack of sufficient statistical power to detect all the true signals. In this work, we propose a solution to this, based on a multi-stage hierarchical Bayesian framework that synthesizes information from both patient and model system databases across multiomic levels to <u>improve the identification of novel cancer driver genes and association with drug responses</u>.

### A Bayesian evidence synthesis procedure

Our integrative framework is called <u>BaySyn</u>: a multistage hierarchical <u>Bay</u>esian evidence <u>syn</u>thesis pipeline for analysis of multi-system multiomic data. The first stage identifies <u>cancer driver genes</u> by detecting transcriptomic associations with upstream changes, which are then utilized to <u>inform biomarker association models</u> in the second stage to improve selection. Specifically, the first stage uses additive Gaussian process regression models to detect potential nonlinear associations of gene expression data with corresponding copy number and methylation profiles for both cell line cancer lineages and patient cancer types. To tackle the issue of lower sample size in cell line data, we propose multi-lineage versions of these <u>mechanistic models</u> that can deconvolve lineage and upstream main effects as well as any potential interactions, in addition to single-lineage versions of the same. Evidence synthesized across a common pool of genes from the two sources is then used in a calibrated Bayesian variable selection procedure in the second stage to identify genes having high association with an outcome variable of interest, such as drug response data. Specifically, the evidence quantifications from the mechanistic models are used in these <u>outcome models</u> to upweight the prior probability of selection of different biomarkers in a spike-and-slab prior setting. A conceptual schematic of the procedure is presented in Figure 1, providing a high-level summary of the multi-model system evidence synthesis through the mechanistic models and calibrated biomarker selection via the outcome models. We apply our framework to multiomic CCLE and TCGA datasets from pan-gynecological cancers (breast, ovary, and uterus lineages). Our mechanistic models provide cancer-specific and cross-lineage evidence that implicate several relevant functional genes such as PTPN6 and ERBB2 in the KEGG adherens junction gene set. Furthermore, our outcome model is able to make higher number of discoveries in drug response models than its uncalibrated counterparts under the same thresholds of type I error control, including detection of known lineage-specific biomarker associations such as BCL11A in breast and FGFRL1 in ovarian cancers.

**Fig. 1:**
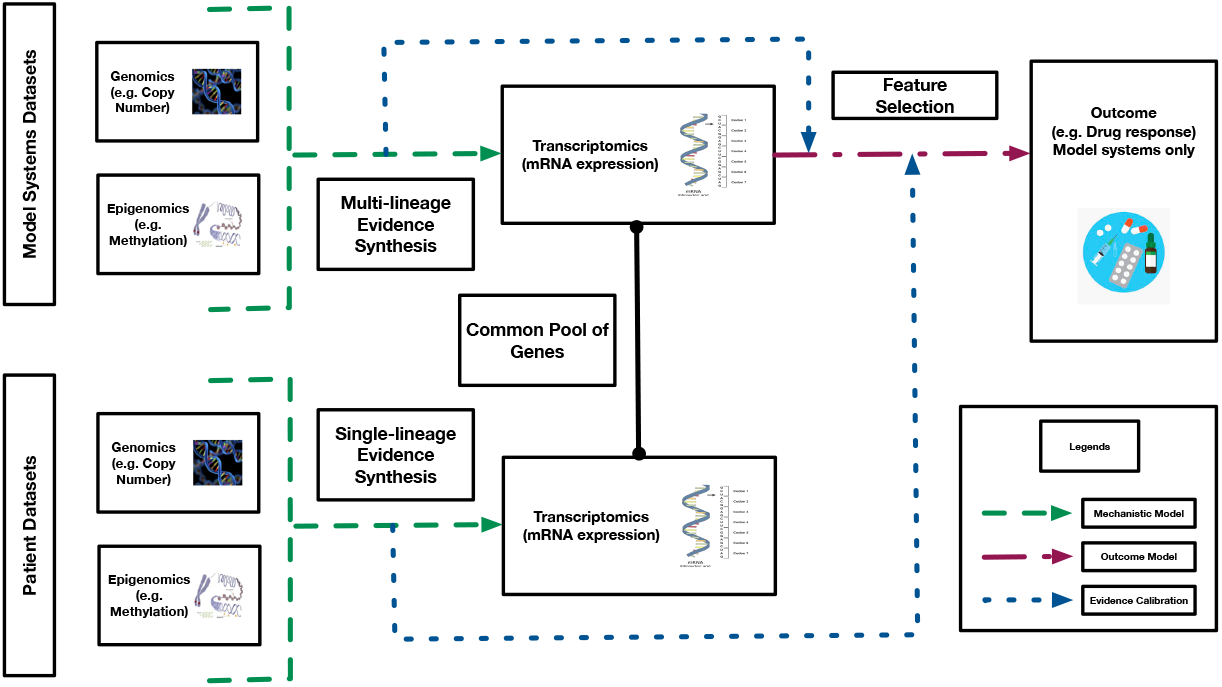
Conceptual schema of the *BaySyn* framework.

The rest of the paper is organized as follows. Section 2 summarizes the multi-stage data integration framework. Section 3 describes the CCLE and TCGA data processing and analysis procedures, along with summarization of interesting results. We finish with a brief discussion of our proposed procedure and findings in Section 4. All the processed datasets, R codes for the pipeline, and the complete set of real data results are available for access via an interactive R Shiny dashboard.

## 2. Methods

### Multi-stage integration pipeline

Following Figure 1, for a given set of samples (patients/model systems), we build gene-specific mechanistic models to infer functional relevance of the genes in the samples of interest based on the association of the gene’s expression pattern with its upstream covariates such as copy number changes or DNA methylation. Particularly, in case of model systems, certain cancer lineages may <u>contain a low number of samples and the mechanistic models may suffer from a lack of sufficient statistical power</u> to identify true associations with upstream factors. Therefore, we build two versions of the mechanistic models depending on the sample size scenarios - a multi-lineage model that can borrow strength across samples from different lineages (used in this work for modeling the cell line samples; Section 2.1.1), and a single-lineage version that can be applied to a set of samples from a single cancer lineage/type (used in this work in context of the patient samples; Section 2.1.2). Based on statistical summaries of significance of the upstream factors for each gene from these models, we then build the outcome-specific Bayesian hierarchical variable selection models (outcome models, in short; Section 2.2) that can incorporate such prior information and borrow strength to improve selection of genes. The specifics of each type of model are described in full detail in the rest of this section.

#### 2.1. Mechanistic Models

For the mechanistic models, we investigate a gene of interest specifically in relation with its upstream factors to detect whether it is a functional driver, and repeat the procedure across the complete pool of genes included in the analyses. This approach offers a highly parallelizable framework, and the efficiency only depends on the <u>computational resources</u> used by each individual model. Further, the class of genomic associations with upstream factors that we are interested in may be highly nonlinear, as has been indicated in past cancer literature.^13,14^ Therefore, we intend to equip our models with sufficiently <u>flexible specifications that can identify a broad range of association patterns</u>. Keeping these useful features in mind, we describe the mathematical details of the multi- and single-lineage mechanistic models below.

##### 2.1.1. Multi-lineage Mechanistic Models

###### Notations

We begin with setting up some notations. Let *M* denote the number of lineages across which we intend to borrow strength in a single mechanistic model, and let {*n*_1_, …, *n*_*M*_ } denote the lineage-specific sample sizes, with 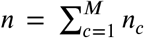 being the total sample size. Across a total of *j* ∈ {1, …, *q*} genes, let *G*_*ij*_ denote the (continuous) normalized expression data for the *j*^th^ gene in the *i*^th^ sample. Let *L*_*i*_denote the lineage (tissue/cancer type) of the *i*^th^ sample, and let **U**_*ij*_ = (*U*_*ij*1_, …, *U*_*ijp*_)^*T*^ *p*_*j*_ × 1 vector of upstream information from sample *i* matched to gene *j*. Our mechanistic models are gene-specific, allowing different sample sizes for each gene. However, for simplicity of notations, we describe the models assuming a fixed *n*.

###### Model structure

For the *j*^th^ gene, we build an additive multi-lineage mechanistic model containing separable components for the main effects of lineage and each upstream covariate, along with any possible interactions of lineage with the upstream factors. Assuming the *G*_*ij*_s to be mean-centered, the general mathematical form of such a model is presented in the following equation.

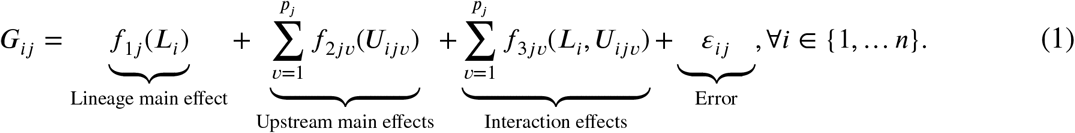

The simplest choice is to specify each component *f* as a linear model. Such models have been explored in context of cancer omics.^15^ Although they are computationally simple, they may not be fully able to capture the general range of cellular association patterns. An obvious nonlinear extension is to use splines to construct piece-wise linear mean profiles. Such approaches have also been explored in this context.^16^ However, there are multifold challenges – including specifying the number of knots (hence the degree of adaptable nonlinearity) and increasing computational intensity with increasing number of covariates. To build a general class of additive association models while maintaining a reasonable extent of computational efficiency, we use Gaussian process (GP) models.

To build an additive GP model with interaction effects, we adapt an existing approach proposed in context of longitudinal data.^17^ In a repeated measures setting, this approach provides a way to incorporate sample-level baseline effects and treatment effects in a nonlinear fashion. We extend this idea to our scenario to include <u>lineage-level baseline effects</u> (treating the experiments on cell lines from the same lineage akin to a repeated experiment setting) and <u>changes in the effects of upstream covariates across different lineages</u>. While samples belonging to cancers sharing some larger group-specific commonalities (e.g. all gynecological cancers) may share patterns of mechanistic impacts of upstream platforms on gene expressions, there may still be cancer-specific differences in the exact effects. Briefly, we use a GP equipped with a zero-sum (zs) kernel for the main effect of the categorical lineage variable, one with an exponentiated quadratic (eq) kernel for the main effects of the continuous upstream variables, and a product of the zs and eq kernels for their interactions, following existing approaches.^17,18^ The specifics of the GP model along with the prior choices are described in detail in [Supplementary Notes Section S1.1](https://drive.google.com/file/d/166rGXVPlqB9g48EP1W7XvKvUrK6ImpWD/view).

###### Model fitting and hypothesis testing

The interest now is in building mechanistic models and testing for different main and interaction effects of interest. We use a dynamic Hamiltonian Monte Carlo (HMC) sampler to obtain draws from the posterior distributions of the parameters. Since we are interested in evaluating the roles of lineage, upstream factors, and any possible interactions in explaining the variability in gene expressions, we are interested in testing the following hypotheses.

1. **Lineage main effect:** *H*_0*Lj*_ : *f*_1*j*_ = constant.
2. **Upstream main effects:** *H*_0*Uj*_ : *f*_2*jν*_ = constant, ∀*ν* ∈ {1, …, *p*_*j*_}.
3. **All upstream effects:** *H*_0*UIj*_ : *f*_2*jν*_, *f*_3*jν*_ = constant, ∀*ν* ∈ {1, …, *p*_*j*_}.

To perform these tests, we use model comparison procedures using HMC-based draws of the joint log-posterior function of the parameters in a model. For a model M containing all or some of the components in Equation (1), let *H*_0·_ be the test of interest and M_·_ be the null model, which is a submodel of M not containing the components set to constant under *H*_0·_. For example, if we are interested in testing the lineage main effect in a main effects-only model M, M_·_ would be an upstream-only model. We define *pseudo-Bayes factors* (pBF_·*j*_s) as scalar summaries of component significance, defined to be the mean difference of the log-posteriors evaluated across the MCMC draws between the two models being compared. The pBFs for the three hypotheses above and for the *j*^th^ gene are denoted respectively by pBF_*Lj*_, pBF_*Uj*_, and pBF_*UIj*_. <u>Note that these quantities are approximations for the traditional log-Bayes factors (lBFs) for comparing Bayesian models under equal model priors.</u> To compute an lBF, one has to compute the expected posteriors for each model, followed by their logratio. Here, we are computing an empirical average of the difference of log-posteriors of the model parameters. The exact expressions of these quantities for a given HMC sample of the parameters are derived in [Supplementary Notes Section S1.2](https://drive.google.com/file/d/166rGXVPlqB9g48EP1W7XvKvUrK6ImpWD/view). We use standard cut-offs for significance used for lBFs at a log_10_(·)-scale: < 0.5 (no evidence), 0.5 − 1 (substantial), 1 − 2 (strong), and > 2 (decisive).^19^ From now on, by pBF we always mean a quantity already in this scale.

###### Sequential evidence detection

To identify driver genes, we quantify evidence of any <u>upstream effect on gene expression untangled from any possible lineage effect</u>. To this end, mimicking classical approaches in regression settings, we follow a sequential scheme as described in [Supplementary Figure S1](https://drive.google.com/file/d/166rGXVPlqB9g48EP1W7XvKvUrK6ImpWD/view).

1. Test for any lineage main effect using pBF_*Lj*_. If pBF_*Lj*_ ≤ 1, go to Step 2. Else go to Step 3.
2. Test *H*_0*U j*_ using pBF_*U j*_. Set mechanistic evidence *ε*_*j*1_ = pBF_*U j*_.
3. Test *H*_0*Uj*_ using pBF_*UI j*_. Set mechanistic evidence *ε*_*j*1_ = pBF_*UI j*_.

##### 2.1.2. Single-lineage Mechanistic Models

These models do not include any lineage main or interaction effects. Thus, from Equation (<u>1</u>), the full models reduce to the following for the *j*^th^ gene, using same notations as before.

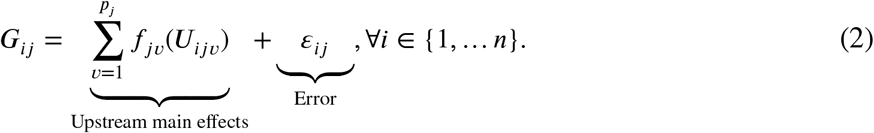

We use the same eq kernel parametrization for the GP priors on each *f* as we used for the *f*_2·_ components in the multi-lineage models. We now test *H*_0*j*_ : *f*_*jν*_ = constant, ∀*ν* ∈ {1, …, *p*_*j*_} for each gene. We compare the full model in Equation (2) with a noise-only null model. The derivation of the corresponding pBF_*j*_ is described in [Supplementary Notes Section S1.3](https://drive.google.com/file/d/166rGXVPlqB9g48EP1W7XvKvUrK6ImpWD/view). We assign the evidence *ε*_*j*2_ = pBF_*j*_, as described in [Supplementary Figure S1](https://drive.google.com/file/d/166rGXVPlqB9g48EP1W7XvKvUrK6ImpWD/view).

#### 2.2. Outcome Model

For a given pool of genes, it is possible to compute multiple lines of evidence (*ε*_*j*_ = (*ε*_*j*1_, …, *ε*_*jE*_)^*T*^ for gene *j*). For example, for a given gene, we may compute <u>one pBF from a multi-lineage model built on cell line samples, and another pBF from a single-lineage model built on patient samples (*E* = 2)</u>. With interest in some disease- or therapy-related phenotype/outcome *Y* and the selection of biomarkers associated with it, the goal is to <u>inform the outcome model about any level of evidence captured in these *ε*_*je*_ s in a covariate-specific way</u> to possibly improve selection.

1. Sufficiently strong evidence in favor of a covariate ⟹ higher prior probability of inclusion.
2. Otherwise, a uniform prior is placed on selection/non-selection for that particular covariate. We utilize a hierarchical Bayesian setting with calibrated spike-and-slab priors, described below. Let *Y*_*i*_ be the mean-centered continuous outcome for the *i*^th^ sample. Simple extensions to categorical/censored outcomes are possible, but in this work we only focus on continuous outcomes. The mathematical form of the calibrated Bayesian variable selection (cBVS) model is then the following.

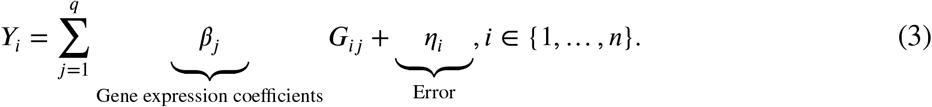

### Model and prior specifications

The errors *η*_*i*_ are iid N(0, *τ*^2^), ∀*i* ∈ {1, …, *n*}. A standard conjugate prior is used for *τ*^2^ ∼ Inverse-Gamma 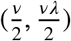. Let ***β*** = (*β*_1_, …, *β*_*q*_)^*T*^ denote the *q*-dimensional vector of regression coefficients. We place a cali^2^bra^2^ted hierarchical spike-and-slab prior on ***β***.

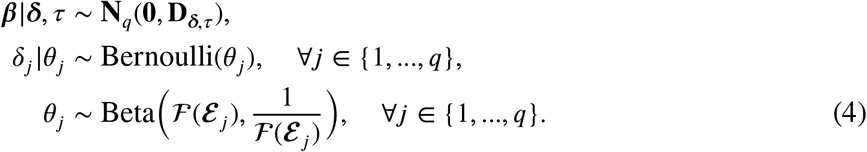

Here **D**_***δ***,*τ*_ = *τ*^2^**A**_***δ***_, where **A**_***δ***_ is the *q*×*q* diagonal matrix **A**_***δ***_ = diag{*δ*_1_*ν*_1_ +(1−*δ*_1_)*ν*_0_, …, *δ*_*q*_*ν*_1_ +(1−*δ*_*q*_)*ν*_0_} and *ν*_1_ ≥ *ν*_0_ > 0 are respectively the slab and spike variances. The binary latent variables *δ*_*j*_ are variable inclusion indicators with *δ*_*j*_ = 1 meaning that the *j*^th^ variable is included in the model. *ℱ* is a calibration function mapping the evidence vector *ε*_*j*_ = (*ε*_*j*1_, …, *ε*_*jE*_)^*T*^ to the prior covariate inclusion probability *θ*_*j*_. <u>The advantages of the hierarchical formulation</u> (Equation (4)) coupled with the evidence calibration function *ℱ* <u>are multifold</u>. First, by adapting *ℱ*, our framework allows the user to incorporate other significance quantities (such as p-values) into the final outcome model. Any external upstream information, including categorical and continuous covariates, can be used in the mechanistic layer to compute such summary statistics. Finally, by tuning *ℱ* appropriately, our framework allows the user to control the impact of the prior information on selection, as we show below. We discuss all these in more detail in Section 4.

### Choice of evidence calibration function

We use a calibration function *ℱ* on ℝ^*E*^ → [0, 1] to aggregate multi-dimensional prior evidence into a scalar prior probability. To this end, we use a four-parameter logistic map reflecting the maximal evidence across all sources on a continuous and nondecreasing spectrum of evidence strength. The exact mathematical form and the motivation behind this choice are described in [Supplementary Notes Section S1.4](https://drive.google.com/file/d/166rGXVPlqB9g48EP1W7XvKvUrK6ImpWD/view). Using this function, the calibrated prior means of *θ*_*j*_ (representative values of maximal evidence at the pBF/ ln(10) scale in parentheses) are as follows: 0.502 (0.25), 0.543 (0.75), 0.726 (1.5), 0.962 (3). As illustrated in [Supplementary Figure S2](https://drive.google.com/file/d/166rGXVPlqB9g48EP1W7XvKvUrK6ImpWD/view), the corresponding prior distributions of *θ*_*j*_ shift from an uniform prior to one concentrated close to one with increase in prior evidence strength.

### Variable selection

Inference is centered around the posterior 𝓅(***β, δ, θ***, *τ*|***Y***, ***G***,, *ν, λ, ν, ν*_0 1_), where ***β, δ***, and ***θ*** are the *q* × 1 vectors of all *β*_*j*_s, *δ*_*j*_s, and *θ*_*j*_s respectively, ***Y*** _*n*×1_ is the outcome vector, ***G***_*n*×*q*_ is the design matrix, and *ε*_*q*×*E*_ is the matrix of the *ε*_*je*_ s. We approximate this using a Gibbs sampler implemented via the *rjags R package*.^20^ We obtain posterior estimates of the parameters 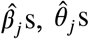 and 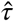 as their corresponding empirical posterior means. Model selection is performed using the collection of 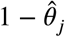 as p-value type quantities and applying a false discovery rate (FDR) control procedure,^21^ described in [Supplementary Notes Section S1.5](https://drive.google.com/file/d/166rGXVPlqB9g48EP1W7XvKvUrK6ImpWD/view).

## 3. Multi-system and Multi-platform Integrative Analyses of Pan-Gynecological Cancers

We perform an integrative analysis of cancer cell lines data from CCLE and patient samples from TCGA.^10,11^ Using multi-lineage mechanistic models for cell line samples and single-lineage mechanistic models for patient samples, we quantify gene-specific associations of expression with corresponding copy number and methylation data. We then use the pBFs from these two sources to inform and build cBVS models of <u>drug response</u> on gene expression based on the cell line samples. Specifically, our multi-lineage mechanistic models on the cell line samples borrow strength by combining data across three gynecological lineages <u>breast, ovary, and uterus</u>. The single-lineage mechanistic models on the patient samples are built separately for each of the three corresponding TCGA cancer types by tissue <u>breast invasive carcinoma (BRCA), ovarian serous cystadenocarcinoma (OV), and uterine carcinosarcoma (UCS)</u>. The outcome models on the cell line samples are built in a lineagespecific way for a collection of drugs of interest in gynecological cancers. Our investigations are aimed broadly at answering two sets of questions.

1. We assess within-system and between-system patterns of functional evidence garnered by the mechanistic models (i.e., a gene may have strong mechanistic evidence of association with the upstream factors for the cell lines only, the patients only, both, or none).
2. We identify panels of genes whose expressions are associated with responses to specific drugs in the cell line samples, potentially offering novel introspection into treatment selection and the cellular mechanisms/targets of such drugs.

### 3.1. Data Processing and Analysis Pipeline

#### Multi-omics cell line and patient data

Gene expression, copy number, and DNA methylation data on cancer cell lines from CCLE, drug response data from GDSC, along with annotation information to match genes to upstream information, are downloaded from the depmap portal.^22^ Gene expression, copy number, and DNA methylation data on TCGA patient samples, along with annotation information matching genes to upstream covariates, are downloaded from the Xena browser.^23^ Sample size and other filtering requirements result in a pool of 5,792 genes and 65 drugs to be included in all further analyses, as described in [Supplementary Notes Section S1.6](https://drive.google.com/file/d/166rGXVPlqB9g48EP1W7XvKvUrK6ImpWD/view). Summary information on each dataset are available in [Supplementary Table S1](https://drive.google.com/file/d/166rGXVPlqB9g48EP1W7XvKvUrK6ImpWD/view) and [Supplementary Figures S3-S8](https://drive.google.com/file/d/166rGXVPlqB9g48EP1W7XvKvUrK6ImpWD/view).

#### BaySyn analysis of gynecological cancers

For each gene, a multi-lineage mechanistic model with *M* = 3 (breast, ovary, uterus) is constructed (termed the CL model hereafter) and hypothesis tests are performed as described in Supplementary Figure S1. Further, for each gene, three single-lineage mechanistic models (one for each cancer type – BRCA, OV, UCS) are built on the patient samples and upstream effects are quantified following [Supplementary Figure S1](https://drive.google.com/file/d/166rGXVPlqB9g48EP1W7XvKvUrK6ImpWD/view). As a post-model fitting investigation, we perform gene set enrichment analyses (GSEA)^24^ using these four sets of evidence (CL, BRCA, UCS, OV) for the Kyoto Encyclopedia of Genes and Genomes (KEGG)^25^ and gene ontology (GO) gene sets.^26,27^ For our analyses, we use the gene set enrichment (GAGE) procedure implemented in the *gage R package* due to the reason that our pBFs are on a different scale than typical expression levels or fold-change summaries.^28^ The gene set-specific hypothesis that we test is whether the set in question exhibits significantly higher level of activity as summarized by the evidence statistics compared to the genes outside the gene set, due to the unidirectional nature of the pBFs. For each lineage, drug-specific response association models are built using the cBVS procedure, and variable selection is performed using a 10% FDR control threshold.

### 3.2. Results

#### Utility of borrowing strength to detect mechanistic evidence

Figure 2a summarizes the number of genes inferred to be at the decisive level of evidence (in favor of associations with corresponding upstream covariates) across the three single-lineage models for each TCGA patient cancer type and the multi-lineage model for the cell lines data. The connected dots at the bottom indicate the intersection of the mechanistic models for which the number of genes summarized by the bar height are decisive. The top three combinations of models in terms of detecting decisive evidence all belong to some combination of the TCGA data sets (BRCA only, BRCA and OV, BRCA and UCS in decreasing order). However, except for the BRCA dataset which utilizes > 750 samples for all genes to build the mechanistic models, the cell lines mechanistic models borrowing strength across three lineages detect more unique signals (4^th^ in the ranking) than the other TCGA datasets. This further validates the utility of building joint nonlinear association models with main and interaction components that can identify shared patterns of association across smaller datasets which would potentially be missed in dataset-specific models. The list of genes uniquely identified by the cell lines mechanistic model is available in [Supplementary Table S2](https://drive.google.com/file/d/166rGXVPlqB9g48EP1W7XvKvUrK6ImpWD/view).

**Fig. 2:**
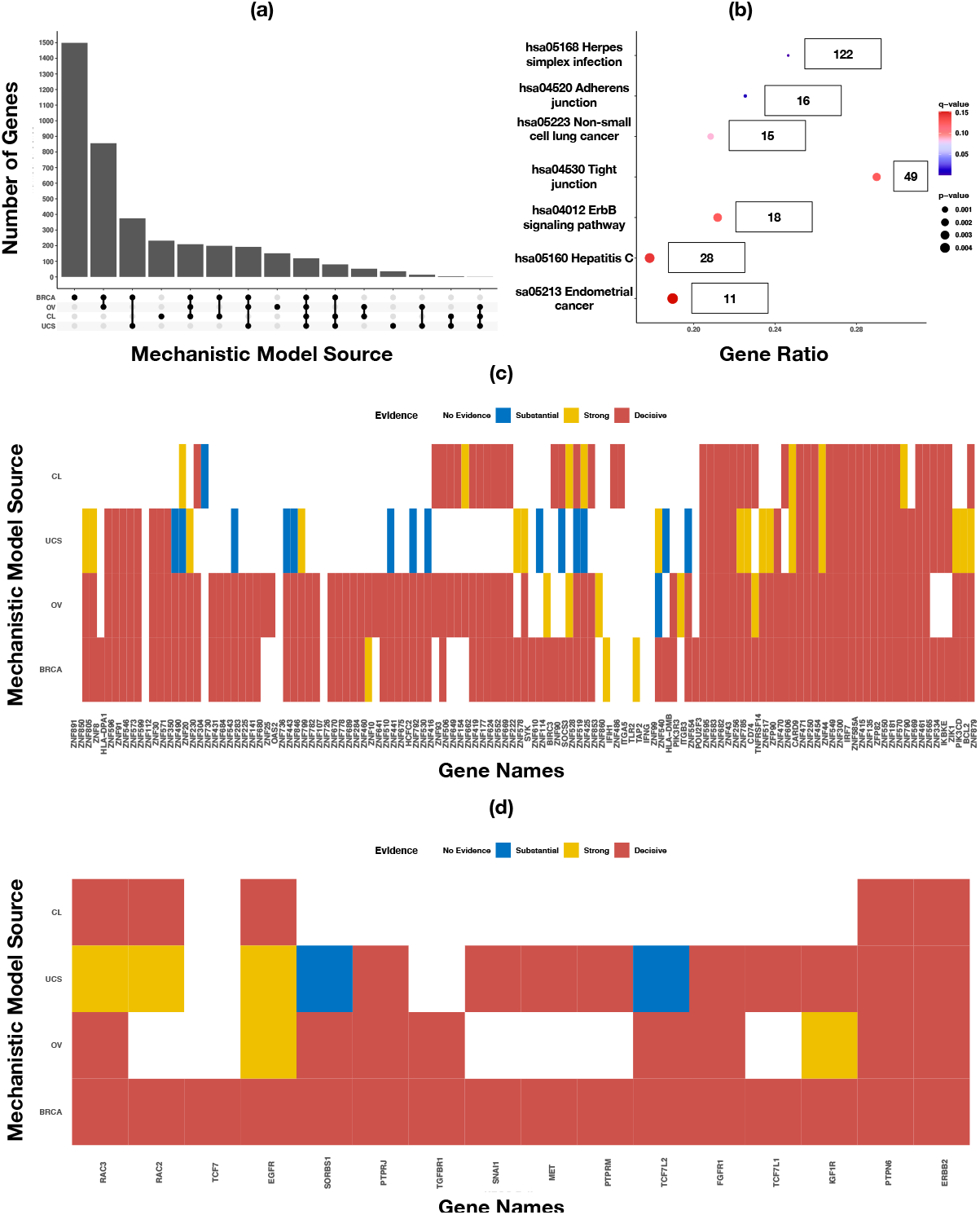
Mechanistic evidence summary and gene set enrichment results. Panel (a) presents an upset plot of the number of genes at the decisive level of evidence based on the mechanistic models for different intersections of the patient and cell line datasets. Panel (b) presents a dotplot summarizing significance levels for KEGG gene sets. The gene sets are ordered from top to bottom in decreasing order of q-values (≤ 0.2 included). The labels beside the dots indicate set sizes in our analyses. Panels (c) and (d) present heatmaps summarizing levels of mechanistic evidence for the genes in KEGG herpes simplex infection and adherens junction gene sets respectively. Genes in the rows are ordered based on clusters resulting from the evidence statistics.

#### KEGG gene set enrichment analyses illustrate utility of mechanistic evidences

To assess the utility of the mechanistic evidence quantities and to validate their use in future detection of novel functional drivers, we perform GSEA using the four evidence sources and the KEGG and GO gene sets. Due to space limitations we only discuss the KEGG results here. The GO results are presented in [Supplementary Figures S14-S29](https://drive.google.com/file/d/166rGXVPlqB9g48EP1W7XvKvUrK6ImpW). Several KEGG gene sets have been implicated to have significant roles generally in cancer^29,30^ and specifically in gynecological cancers.^31–34^ The results from our KEGG GSEA are summarized in Figure 2b, exhibiting the seven gene sets with FDR-controlled q-value < 0.2. The gene set-specific mechanistic evidences are summarized in Figure 2c-d for the top two KEGG gene sets; the rest are presented in [Supplementary Figures S9-S13](https://drive.google.com/file/d/166rGXVPlqB9g48EP1W7XvKvUrK6ImpWD/view). The top gene set identified in the KEGG analyses is the herpes simplex infection pathway (p-value = 3.88 × 10^−16^) (Figure 2b). This gene set contains a large cluster of genes exhibiting decisive evidence across majority of the mechanistic models, as can be seen in Figure 2c. Following these genes are two major clusters one containing genes at the decisive level for the BRCA, OV, and CL mechanistic models, and one containing genes at the decisive level for all three TCGA cancers. The consistent nature of functional evidence across this gene set is in agreement with findings from past investigations - multiple studies have indicated the prognostic value of members of this pathway in gynecological cancers - including breast,^35^ ovarian,^36^ and endometrial^37^ cancer. The second-highest gene set in the KEGG analyses is the adherens junction gene set (p-value = 5.52 × 10^−5^) (Figure 2b). The genes PTPN6 and ERBB2 exhibit decisive levels of mechanistic evidence in all four models (Figure 2d). Different upstream mechanisms of the ERBB2 gene have been implicated in different gynecological cancers, such as copy number changes in ovarian tumors^38^ and somatic mutations in breast cancer.^39^ The EGFR gene has also shown promise as a potential therapeutic target in multiple gynecological cancers,^40,41^ which is in alignment with our findings of some signal in all the TCGA and cell line models (Figure 2d).

#### Calibrated drug response models identify high-association lineage-specific biomarkers

We build calibrated hierarchical Bayesian variable selection-based drug response models for each lineage × drug combination across all 65 drugs and all three cell line lineages. Figure 3a presents a wordcloud where each gene is weighted by the total number of times it is selected in a drug response model at the 10% FDR-controlled cutoff. The genes BAHCC1, ALOX12P2, and SYCP2 emerge as the top candidates, with selection in 14, 12, and 12 models respectively. While this summary allows us to identify general candidates for future pharmacogenomic investigations, it does not indicate any potential lineage-specific utility of these genes. To this end, Figure 3b summarizes the number of times the top genes across all drug response models are selected in each lineage. For breast, genes BAHCC1, BCL11A, and SYCP2 are at the top, with respectively eight, eight, and six detected drug associations. <u>The role of BCL11A in triple-negative breast cancer (TNBC) stemness is well known, and it is considered to be one of the first utilizable targets for treatment of TNBCs</u>.^42^ A similar confirmation can be obtained for SYCP2, which has recently been identified as a prognostic biomarker in breast cancer.^43^ However, to the best of our knowledge, BAHCC1 has not so far been identified to have breast cancer-specific functional roles, which renders it as a novel detection that deserves deeper investigations. Top genes in the two other lineages also include both novel and known functional drivers - such as ALOX12P2 (nine selections, novel) and FGFRL1 (eight selections, known)^44^ for ovary and FBXO17 (seven selections, novel) for uterus.

**Fig. 3:**
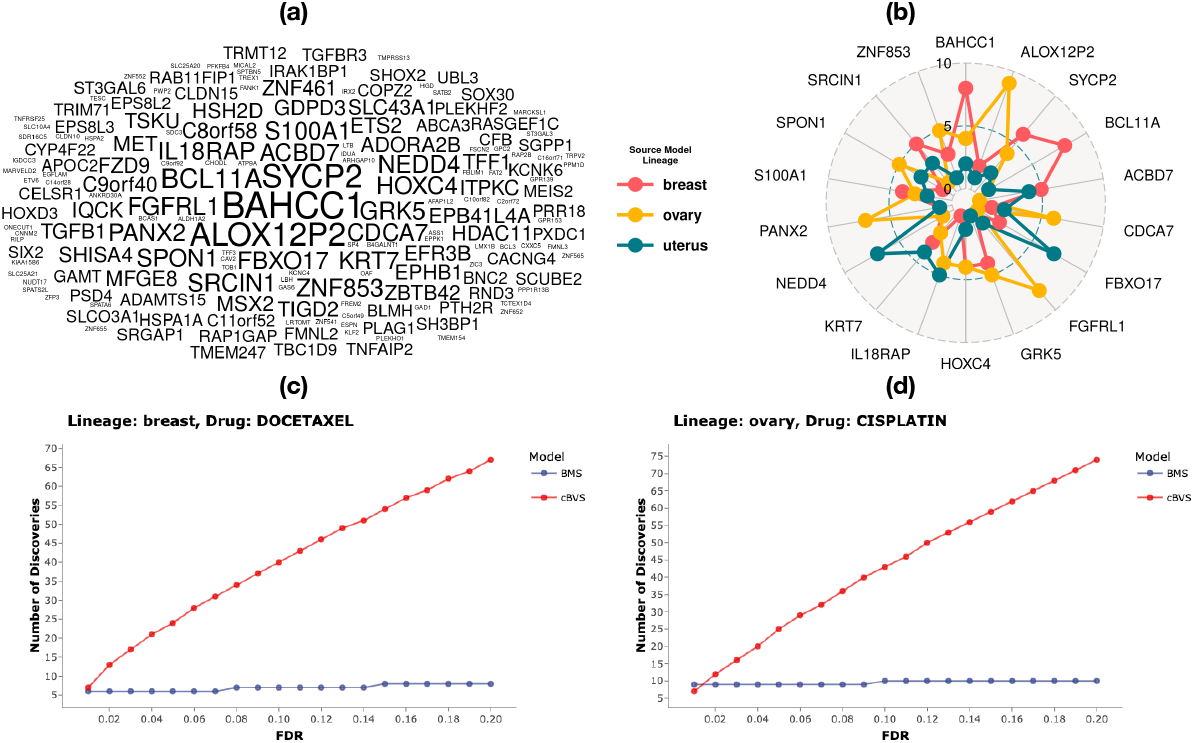
Drug response model summaries. Panel (a) presents a wordcloud of top genes across all the drug response models (for three lineages and 65 drugs). The sizes of the words are proportional to the total number of times across all models that a gene is selected based on a 10% FDR-controlled threshold. Panel (b) presents a radar chart of the top 18 genes (selected in at least nine out of all the drug response models) according to the three lineages. Panels (c) and (d) present discovery plots across increasing FDR control thresholds for the drug docetaxel in lineage breast and the drug cisplatin in lineage ovary respectively. BMS refers to an uncalibrated Bayesian variable selection model based on the Bayesian model averaging procedure (see [Supplementary Notes Section S1.7](https://drive.google.com/file/d/166rGXVPlqB9g48EP1W7XvKvUrK6ImpWD/view)).

#### Calibration improves statistical power to detect gene-drug associations

To assess the discoveries for specific lineage × drug combinations, we focus on two drugs with known use in specific cancer lineages - docetaxel for breast and cisplatin for ovary. The number of discoveries across different FDR thresholds for these two combinations are presented in Figure 3c-d and the corresponding discoveries are summarized in [Supplementary Tables S3-S4](https://drive.google.com/file/d/166rGXVPlqB9g48EP1W7XvKvUrK6ImpWD/view). Similar plots and tables for all other models are available in our R Shiny dashboard. Evidently, compared to an uncalibrated Bayesian variable selection procedure implemented via the BMS R package (see [Supplementary Notes Section S1.7](https://drive.google.com/file/d/166rGXVPlqB9g48EP1W7XvKvUrK6ImpWD/view)), cBVS models make more discoveries at the same level of error control, allowing a continuum of assessment for top candidates emerging across increasing control thresholds. This indicates the utility of synthesizing mechanistic evidence and calibrating the outcome models with such evidences. Several examples of cell lines-based discoveries guided by evidences discovered in patient data emerge. For example, the model for docetaxel response in breast cell lines identify an association with the gene GRK5 at 10% FDR control. Cell lines overexpressing GRK5 have previously been observed to demonstrate an increase in resistance to docetaxel in male gynecological cancers,^45^ and our finding suggests that it deserves further investigations in female gynecological cancers as well. Another top discovery at the same FDR threshold is the gene CD83, expression of which is known to be enhanced by docetaxel in metastatic breast cancers.^46^ For the response model of cisplatin in the ovarian lineage, multiple solute-carrier family (SLC) genes are selected at the 10% FDR threshold. These genes are known potential biomarkers of ovarian cancer and are under investigation for prognostic utility.^47^ Another interesting discovery is that of the CDCA7 gene from the cell division cycle pathway, silencing of which has recently been shown to downregulate cisplatin resistance in lung cancer subtypes, making it a potential therapeutic target.^48^ Our finding seems to indicate similar scope in ovarian cancer, demanding further investigation. Notably, <u>all four of these discussed findings had no cell lines-based mechanistic evidence, but had decisive evidence from at least one TCGA source</u> – which further underscores the importance of synthesizing evidence across model systems.

## 4. Summary and Discussion

We propose BaySyn, a hierarchical multi-stage Bayesian evidence synthesis procedure for multisystem multiomic integration. BaySyn detects functionally relevant driver genes based on their associations with upstream regulators and uses this information to guide variable selection in outcome association models. We apply our framework to multiomic cancer cell line and patient datasets for pan-gynecological cancers. pBFs from the mechanistic layer of BaySyn exhibit high enrichment in previously known KEGG gene sets and detect driver genes known to have functional roles in the cancers studied. Calibrated outcome models for drug responses identify several confirmatory and novel lineage-drug-gene combinations providing further evidence on the profitability of our approach towards future precision oncology endeavors.

Several features of our framework makes it readily adaptable to more general settings and richer datasets. The calibrated spike-and-slab prior can be generalized to include any number (more upstream platforms such as miRNA or mutation) and form (other evidence metrics such as p-values) of prior information by tuning the calibration function accordingly. The outcome model can easily be extended to include other biomarkers such as proteomics. While we use cell lines data to illustrate the integrative approach across model systems, it is straightforward to apply our pipeline to datasets from cancer model systems with higher fidelity to human tumors^49^ - such as organoids^50^ or patient-derived xenografts^51^ - as such databases become increasingly comprehensive and available. Further, both the stages of our framework are highly parallelizable and individual runs are quite efficient - a single gene-specific multi-lineage mechanistic model with interactions takes approximately 20 minutes on average to complete, while a single lineage-drug specific outcome model takes approximately 12 minutes on average (both based on runs on a single core of a 2015 Macbook Air with 8 GB memory and Intel i5 processor). Thus, extending our analyses to include larger gene-drug panels with similar sample sizes is straightforward with existing parallel computing resources.

Certain improvements are of interest given the biological context of our work. First, although we assess mechanistic relevance at a gene-by-gene basis, at a molecular level, genes interact in functional pathways to result in downstream modifications or aberrations. This motivates joint models for driver genes in a multivariable mechanistic model setting that can account for underlying genegene interactions – which requires non-trivial expansions of our model. Second, the relatively low lineage-specific sample sizes in the outcome models in this work make HMC-based fully Bayesian exploration of the posteriors feasible. Increasing sample sizes could result in increased computation times especially for the outcome models; where-in approximate Bayesian computation schemes such as the E-M based variable selection^52^ or variational Bayes^53^ could be employed. We leave these tasks for future exploration.

## Data Availability

All the processed datasets, R codes for the pipeline, and the complete set of real data results are available for access via an interactive R Shiny dashboard at https://rupamb.shinyapps.io/BaySyn/.

