## Supplementary Materials for "BaySyn: Bayesian Evidence Synthesis for Multi-system Multiomic Integration"

### S1. Supplementary Notes

#### S1.1. *Additive Gaussian Process Specification for Multi-lineage Mechanistic Models*

To build an additive GP model with interaction effects, we adapt an existing approach proposed in the context of longitudinal data.[1] In a repeated measures setting, this approach provides a way to incorporate sample-level baseline effects and treatment effects in a nonlinear fashion. We extend this idea to our scenario to include lineage-level baseline effects (treating the experiments on cell lines from the same lineage akin to a repeated experiment setting) and changes in the effects of upstream covariates across different lineages. While samples belonging to cancers sharing some larger group-specific commonalities (e.g. all gynecological cancers) may share patterns of mechanistic impacts of upstream platforms on gene expressions, there may still be cancer-specific differences in the exact effects. We now specify the distributions of the different components in Equation 1 in the main manuscript.

**GP specification and priors** We build a Gaussian likelihood by first assuming $\epsilon_{ij} \overset{\text{iid}}{\sim} \mathrm{N}(0, \sigma_j^2)$, and we then build the $f_{\bullet}$ components using GP priors for each component. For each component $f_{\bullet}$, let us define $f^{(\bullet)} := [f_{\bullet}(\mathbf{x}_1), \ldots, f_{\bullet}(\mathbf{x}_n)]^T$ where $\mathbf{x}_i$ generally denotes the vector of all possible covariates for sample $i$. We assume that this has a multivariate normal prior as $f^{(\bullet)} \sim \mathbf{N}_n(\mathbf{0}, \mathbf{K}^{(\bullet)})$ where the $n \times n$ covariance matrix has entries $\mathbf{K}_{ih}^{(\bullet)} = \alpha_{\bullet}^2 k_{\bullet}(\mathbf{x}_i, \mathbf{x}_h)$. There are a few things worth noting about this model. First, the components $f_{\bullet}$ are assumed independent *a priori*, hence their sum is also a zero-mean GP with kernel $k(\mathbf{x}, \mathbf{x}') = \sum_{\bullet} \alpha_{\bullet}^2 k_{\bullet}(\mathbf{x}, \mathbf{x}')$.[2] Second, each $\alpha_{\bullet}^2$ controls the marginal variance of the corresponding component, while the base kernel function $k_{\bullet}$ controls the component's shape and the induced covariance structure. Third, in our applications, kernels $k_{1j}$ and $k_{2jv}$s are functions of the lineage and upstream covariates only, respectively, while the kernels $k_{3jv}$s are functions of all available covariates. The final step in the model building is to choose the specific kernel functions for each component according to the types and scales of the covariates they take as inputs, as described below.

(1) For component 1 (one categorical covariate), we use the zero-sum (zs) kernel. The marginal variance is denoted by $\alpha_{1j}^2$. Assuming that the model includes samples from a total of $M$ lineages,

© 2022 The Authors. Open Access chapter published by World Scientific Publishing Company and distributed under the terms of the Creative Commons Attribution Non-Commercial (CC BY-NC) 4.0 License.

the kernel is defined as $k_{1j}(L_i, L_h) = \begin{cases} 1 & \text{if } L_i = L_h, \\ \frac{1}{1-M} & \text{else.} \end{cases}$
Note that this choice of kernel function is equivalent to assuming the lineage effects follow a standard random effects model with a zero-sum constraint on the random effects. Namely, this is equivalent to assuming that $f_{1j}(L) \sim N(0, \alpha^2_{1j})$ for $L = 1, \ldots, M$ independently, with the constraint that $\sum_{L=1}^{M} f_{1j}(L) = 0$.

(2) For component 2 (only continuous upstream covariates), we use the exponentiated quadratic (eq) kernel on each covariate. The kernel for the $v^{\text{th}}$ upstream covariate corresponding to gene $j$ is defined as $k_{2jv}(U_{ijv}, U_{hjv}) = \exp(-\frac{(U_{ijv}-U_{hjv})^2}{2l^2_{2jv}})$. The $j^{\text{th}}$ marginal variance is denoted by $\alpha^2_{2jv}$.

(3) For component 3 (interactions between categorical lineage information and continuous upstream covariates), we use the product of zs and eq kernels on each interaction. For the $v^{\text{th}}$ interaction, the kernel is defined as $k_{3jv}((L_i, U_{ijv})^T, (L_h, U_{hjv})^T) = k_{1j}(L_i, L_h)\exp(-\frac{(U_{ijv}-U_{hjv})^2}{2l^2_{3jv}})$ following existing approaches.[3] The $j^{\text{th}}$ marginal variance is denoted by $\alpha^2_{3jv}$.

Each marginal standard deviation $\alpha_{\bullet}$ is given a Student-$t^{+}_{20}$ prior, and each length-scale parameter $l_{\bullet}$ is given a Log-Normal(0, 1) prior, independently. The residual variance paramters $\sigma^2_j$ are assigned an Inverse-Gamma(2, 1) prior.

#### S1.2. *Hypothesis Testing for Multi-lineage Mechanistic Models*

**Model fitting** The interest is in building mechanistic models that would allow us to test for different main and interaction effects of interest. Due to the nature of the zs kernel, the interaction components will also have the zero-sum property,[1] which makes it simple to extract and interpret the interaction effects separately. We use a dynamic Hamiltonian Monte Carlo (HMC) sampler as implemented in the *R package lgpr* to obtain draws from the posterior distributions of the parameters, and arrive at the posterior of the functional components analytically for Gaussian likelihood.[1] While selection of the specific components and hence covariates is possible based on ranking Bayesian variable relevance statistics or following a minimal subset selection-type approach using such statistics, we are more interested in quantifying the significance of the main and interaction effects as separate collectives, and follow the approach described below.

**Model comparison and testing** Since we are interested in evaluating the roles of lineage, upstream factors, and any possible interactions in explaining the variability in gene expressions, we are interested in testing the following hypotheses for the $j^{\text{th}}$ gene.

(1) **Lineage main effect:** $H_{0Lj} : f_{1j} = \text{constant}$.
(2) **Upstream main effects:** $H_{0Uj} : f_{2jv} = \text{constant}, \forall v \in \{1, \ldots, p_j\}$.
(3) **All upstream effects:** $H_{0UIj} : f_{2jv}, f_{3jv} = \text{constant}, \forall v \in \{1, \ldots, p_j\}$.

To perform these tests, we need to be able to construct models that contain the additive components of interest and compare them against submodels without those components. We use log-posteriors of the parameters in a model to perform the model comparisons, computing HMC-based pseudo-Bayes factors (pBF$_{\bullet j}$s) as scalar summaries of component significance. First, we describe the models we construct and the log-posterior (LP) quantities for each below. Here $\mathbf{G}_{.j} = (G_{1j}, \ldots, G_{nj})^T$.

(M1) **Lineage-only model:** Components $f_{2\bullet}$ and $f_{3\bullet}$ in Equation 1 in the main manuscript are not included. The expression for the log-posterior of this model is given below. Here $A = -\frac{n}{2}\ln(2\pi)$ and $B = \ln\frac{2\Gamma(10.5)}{\sqrt{20\pi}\Gamma(10)}$ are constants free of the model parameters and data. $\boldsymbol{\Sigma}_{1j} = \mathbf{V}_{0j} + \mathbf{V}_{1j}$ where $\mathbf{V}_{0j} = \sigma_j^2 \mathbf{I}_n$ and $\mathbf{V}_{1jih} = \alpha_{1j}^2 \mathscr{k}_{1j}(L_i, L_h)$.

$$LP_{1j} = \ln[\mathscr{p}(\mathbf{G}_{\bullet j}|\alpha_{1j}, \sigma_j^2).\mathscr{p}(\sigma_j^2).\mathscr{p}(\alpha_{1j})]$$

$$= \ln[(2\pi)^{-\frac{n}{2}}|\boldsymbol{\Sigma}_{1j}|^{-\frac{1}{2}}\exp\{-\frac{1}{2}\mathbf{G}_{\bullet j}^T\boldsymbol{\Sigma}_{1j}^{-1}\mathbf{G}_{\bullet j}\}.\Gamma(2)^{-1}\sigma_j^{-6}\exp(-\sigma_j^{-2}).\frac{2\Gamma(10.5)}{\sqrt{20\pi}\Gamma(10)}(1+\frac{\alpha_{1j}^2}{20})^{-10.5}I(\alpha_{1j}>0)]$$

$$= A + B - \frac{\ln|\boldsymbol{\Sigma}_{1j}| + \mathbf{G}_{\bullet j}^T\boldsymbol{\Sigma}_{1j}^{-1}\mathbf{G}_{\bullet j}}{2} - 6\ln\sigma_j - \sigma_j^{-2} - 10.5\ln(1+\frac{\alpha_{1j}^2}{20}) + \ln I(\alpha_{1j}>0). \quad (1)$$

(M2) **Upstream-only model:** Components $f_1$ and $f_{3\bullet}$ in Equation 1 in the main manuscript are not included. The expression for the log-posterior of this model is given below. Here $\boldsymbol{\Sigma}_{2j} = \mathbf{V}_{0j} + \mathbf{V}_{2j}$ where $\mathbf{V}_{2jih} = \sum_{v=1}^{p_j}\alpha_{2jv}^2\mathscr{k}_{2jv}(U_{ijv}, U_{hjv})$.

$$LP_{2j} = \ln[\mathscr{p}(\mathbf{G}_{\bullet j}|\alpha_{2j1},\ldots,\alpha_{2jp_j}, l_{2j1},\ldots,l_{2jp_j},\sigma_j^2).\mathscr{p}(\sigma_j^2).\prod_{v=1}^{p_j}\{\mathscr{p}(\alpha_{2jv})\mathscr{p}(l_{2jv})\}]$$

$$= \ln[(2\pi)^{-\frac{n}{2}}|\boldsymbol{\Sigma}_{2j}|^{-\frac{1}{2}}\exp\{-\frac{1}{2}\mathbf{G}_{\bullet j}^T\boldsymbol{\Sigma}_{2j}^{-1}\mathbf{G}_{\bullet j}\}.\Gamma(2)^{-1}\sigma_j^{-6}\exp(-\sigma_j^{-2}).\prod_{v=1}^{p_j}\{\frac{2\Gamma(10.5)}{\sqrt{20\pi}\Gamma(10)}(1+\frac{\alpha_{2jv}^2}{20})^{-10.5}I(\alpha_{2jv}>0)\}$$

$$.\prod_{v=1}^{p_j}\{\frac{1}{\sqrt{2\pi}l_{2jv}}\exp(-\frac{(\ln(l_{2jv}))^2}{2})\}]$$

$$= \frac{n+p_j}{n}A + p_jB - \frac{\ln|\boldsymbol{\Sigma}_{2j}| + \mathbf{G}_{\bullet j}^T\boldsymbol{\Sigma}_{2j}^{-1}\mathbf{G}_{\bullet j}}{2} - 6\ln\sigma_j - \sigma_j^{-2} - 10.5\sum_{v=1}^{p_j}\ln(1+\frac{\alpha_{2jv}^2}{20}) + \sum_{v=1}^{p_j}\ln I(\alpha_{2jv}>0)$$

$$-\sum_{v=1}^{p_j}\ln(l_{2jv}) - \frac{1}{2}\sum_{v=1}^{p_j}(\ln(l_{2jv}))^2. \quad (2)$$

(M3) **All main effects model:** Components $f_{3\bullet}$ in Equation 1 in the main manuscript are not included. The expression for the log-posterior of this model is given below. Here $\boldsymbol{\Sigma}_{3j} = \mathbf{V}_{0j} + \mathbf{V}_{1j} + \mathbf{V}_{2j}$.

$$LP_{3j} = \frac{n+p_j}{n}A + (p_j+1)B - \frac{\ln|\boldsymbol{\Sigma}_{3j}| + \mathbf{G}_{\bullet j}^T\boldsymbol{\Sigma}_{3j}^{-1}\mathbf{G}_{\bullet j}}{2} - 6\ln\sigma_j - \sigma_j^{-2} - 10.5\ln(1+\frac{\alpha_{1j}^2}{20}) + \ln I(\alpha_{1j}>0)$$

$$-10.5\sum_{v=1}^{p_j}\ln(1+\frac{\alpha_{2jv}^2}{20}) + \sum_{v=1}^{p_j}\ln I(\alpha_{2jv}>0) - \sum_{v=1}^{p_j}\ln(l_{2jv}) - \frac{1}{2}\sum_{v=1}^{p_j}(\ln(l_{2jv}))^2. \quad (3)$$

(M4) **Interactions model:** All components in Equation 1 in the main manuscript are included. The expression for the log-posterior of this model is given below. Here $\boldsymbol{\Sigma}_{4j} = \mathbf{V}_{0j} + \mathbf{V}_{1j} + \mathbf{V}_{2j} + \mathbf{V}_{3j}$ where $\mathbf{V}_{3jih} = \sum_{v=1}^{p_j}\alpha_{3jv}^2\mathscr{k}_{3jv}((L_i, U_{ijv})^T, (L_h, U_{hjv})^T)$.

$$LP_{4j} = \frac{n+2p_j}{n}A + (2p_j+1)B - \frac{\ln|\boldsymbol{\Sigma}_{4j}| + \mathbf{G}_{\bullet j}^T\boldsymbol{\Sigma}_{4j}^{-1}\mathbf{G}_{\bullet j}}{2} - 6\ln\sigma_j - \sigma_j^{-2} - 10.5\ln(1+\frac{\alpha_{1j}^2}{20}) + \ln I(\alpha_{1j}>0)$$

$$-10.5\sum_{v=1}^{p_j}\ln(1+\frac{\alpha_{2jv}^2}{20}) + \sum_{v=1}^{p_j}\ln I(\alpha_{2jv}>0) - 10.5\sum_{v=1}^{p_j}\ln(1+\frac{\alpha_{3jv}^2}{20}) + \sum_{v=1}^{p_j}\ln I(\alpha_{3jv}>0)$$

$$- \sum_{v=1}^{p_j} \ln(l_{2jv}) - \frac{1}{2}\sum_{v=1}^{p_j}(\ln(l_{2jv}))^2 - \sum_{v=1}^{p_j} \ln(l_{3jv}) - \frac{1}{2}\sum_{v=1}^{p_j}(\ln(l_{3jv}))^2. \quad (4)$$

**Sequential evidence detection using pBFs** Based on these quantities we now perform the tests of hypotheses as follows. We focus first on model M3 to test whether the lineage component has any effect at all, and move on to M4 (including interactions) only if the answer to the previous question is yes. For model M3, let $S$ denote the number of draws from the HMC sampler, and let $\boldsymbol{\phi}_j^{(s)} = (\alpha_{1j}^{(s)}, \alpha_{2j1}^{(s)}, \ldots, \alpha_{2jp_j}^{(s)}, l_{2j1}^{(s)}, \ldots, l_{2jp_j}^{(s)}, \sigma_j^{(s)})^T$ denote the vector of sampled parameter values at the $s^{\text{th}}$ iteration, $s \in \{1, \ldots, S\}$. Let $LP_{3j}^{(s)}$ and $LP_{2j}^{(s)}$ denote the values of $LP_{3j}$ and $LP_{2j}$ respectively, evaluated at $\boldsymbol{\phi}_j^{(s)}$.

Let $\text{pBF}_{Lj} = \frac{1}{S}\sum_{s=1}^{S}(LP_{3j}^{(s)} - LP_{2j}^{(s)})$ be defined as the pseudo-Bayes factor for testing $H_{0Lj} : f_{1j} =$ constant (lineage main effect). Note that this quantity is an approximation for the log-Bayes factor (lBF) for comparing models M3 and M2 under equal model priors. To compute the lBF, one has to compute the expected posteriors for each models, followed by taking a ratio of the two quantities, followed by a log. Here, we are computing an empirical average of the difference of log-posteriors of the model parameters based on the HMC samples. We use standard cut-offs for significance used for lBFs at a $\log_{10}(\bullet)$-scale: $< 0.5$ (no evidence), $0.5 - 1$ (substantial), $1 - 2$ (strong), and $> 2$ (decisive).[4] From now on, by pBF we always mean a quantity already in this scale. If this test indicates non-significance (neither strong nor decisive evidence), we test $H_{0Uj} : f_{2jv} = \text{constant}, \forall v \in \{1, \ldots, p_j\}$ (upstream main effects), by comparing M3 with M1 via computing $\text{pBF}_{Uj} = \frac{1}{S}\sum_{s=1}^{S}(LP_{3j}^{(s)} - LP_{1j}^{(s)})/\ln(10)$ following similar notations as before. The mechanistic evidence $\mathcal{E}_{j1}$ is then set equal to $\text{pBF}_{Uj}$.

If $\text{pBF}_{Lj}$ falls in the strong or decisive evidence range, we test $H_{0UIj} : f_{2jv}, f_{3jv} = \text{constant}, \forall v \in \{1, \ldots, p_j\}$ (all upstream effects), comparing M4 with M1. For model M4, following our previous notations, let $S$ again denote the number of draws from the HMC sampler, and let $\boldsymbol{\psi}_j^{(s)} = (\alpha_{1j}^{(s)}, \alpha_{2j1}^{(s)}, \ldots, \alpha_{2jp_j}^{(s)}, \alpha_{3j1}^{(s)}, \ldots, \alpha_{3jp_j}^{(s)}, l_{2j1}^{(s)}, \ldots, l_{2jp_j}^{(s)}, l_{3j1}^{(s)}, \ldots, l_{3jp_j}^{(s)}, \sigma_j^{(s)})^T$ denote the vector of sampled parameter values at the $s^{\text{th}}$ iteration, $s \in \{1, \ldots, S\}$. Let $LP_{4j}^{(s)}$ and $LP_{1j}^{(s)}$ now denote the values of $LP_{4j}$ and $LP_{1j}$ respectively, evaluated at $\boldsymbol{\psi}_j^{(s)}$. Let $\text{pBF}_{UIj} = \frac{1}{S}\sum_{s=1}^{S}(LP_{4j}^{(s)} - LP_{1j}^{(s)})/\ln(10)$ be defined as the pseudo-Bayes factor for all (main + interaction) upstream effects. Based on $\text{pBF}_{UIj}$, we can then: (1) assign a level of significance using the standard cut-offs as before, and (2) assign the mechanistic evidence for gene $j$ as $\mathcal{E}_{j1} = \text{pBF}_{UIj}$. The entire testing procedure is then performed independently for each of the $q$ genes, as described in Figure S1.

### S1.3. *Hypothesis Testing for Single-lineage Mechanistic Models*

We are now interested in evaluating the roles of upstream factors, via testing the hypothesis $H_{0j} : f_{jv} = \text{constant}, \forall v \in \{1, \ldots, p_j\}$ for each gene. To utilize the same approach of as before, we compare the following two models, in reference with Equation 2 in the main manuscript.

(M5) **Null model:** Components $f_\bullet$ in Equation 2 in the main manuscript are not included. The expression for the log-posterior of this model is given below. Here $A$, $\mathbf{G}_{\bullet j}$, and $\mathbf{V}_{0j}$ are as before.

$$\begin{aligned} LP_{5j} &= \ln[p(\mathbf{G}_{\bullet j}|\sigma_j^2).p(\sigma_j^2)] \\ &= \ln[(2\pi)^{-\frac{n}{2}}|\mathbf{V}_{0j}|^{-\frac{1}{2}}\exp\{-\frac{1}{2}\mathbf{G}_{\bullet j}^T\mathbf{V}_{0j}^{-1}\mathbf{G}_{\bullet j}\}.\Gamma(2)^{-1}\sigma_j^{-6}\exp(-\sigma_j^{-2}) \end{aligned}$$

$$= A - \frac{\ln|\mathbf{V}_{0j}| + \mathbf{G}_{\bullet j}^T \mathbf{V}_{0j}^{-1} \mathbf{G}_{\bullet j}}{2} - 6\ln\sigma_j - \sigma_j^{-2}. \tag{5}$$

(M6) **Full model:** Components $f_{\bullet}$ in Equation 2 in the main manuscript are included. The expression for the log-posterior of this model is given below. Here $\mathbf{\Sigma}_{5j} = \mathbf{V}_{0j} + \mathbf{V}_{5j}$ where $\mathbf{V}_{5jih} = \sum_{v=1}^{p_j} \alpha_{jv}^2 \mathscr{k}_{jv}(U_{ijv}, U_{hjv}) = \sum_{v=1}^{p_j} \alpha_{jv}^2 \exp(-\frac{(U_{ijv}-U_{hjv})^2}{2l_{jv}^2})$, and $A$, $B$ are as before.

$$LP_{6j} = \ln[\mathscr{p}(\mathbf{G}_{\bullet j}|\alpha_{j1}, \ldots, \alpha_{jp_j}, l_{j1}, \ldots, l_{jp_j}, \sigma_j^2).\mathscr{p}(\sigma_j^2). \prod_{v=1}^{p_j} \{\mathscr{p}(\alpha_{jv})\mathscr{p}(l_{jv})\}]$$

$$= \ln[(2\pi)^{-\frac{n}{2}} |\mathbf{\Sigma}_{5j}|^{-\frac{1}{2}} \exp\{-\frac{1}{2}\mathbf{G}_{\bullet j}^T \mathbf{\Sigma}_{5j}^{-1} \mathbf{G}_{\bullet j}\}.\Gamma(2)^{-1}\sigma_j^{-6}\exp(-\sigma_j^{-2}). \prod_{v=1}^{p_j} \{\frac{2\Gamma(10.5)}{\sqrt{20\pi}\Gamma(10)}(1 + \frac{\alpha_{jv}^2}{20})^{-10.5} I(\alpha_{jv} > 0)\}$$

$$. \prod_{v=1}^{p_j} \{\frac{1}{\sqrt{2\pi} l_{jv}} \exp(-\frac{(\ln(l_{jv}))^2}{2})\}]$$

$$= \frac{n + p_j}{n} A + p_j B - \frac{\ln|\mathbf{\Sigma}_{5j}| + \mathbf{G}_{\bullet j}^T \mathbf{\Sigma}_{5j}^{-1} \mathbf{G}_{\bullet j}}{2} - 6\ln\sigma_j - \sigma_j^{-2} - 10.5 \sum_{v=1}^{p_j} \ln(1 + \frac{\alpha_{jv}^2}{20}) + \sum_{v=1}^{p_j} \ln I(\alpha_{jv} > 0)$$

$$- \sum_{v=1}^{p_j} \ln(l_{jv}) - \frac{1}{2} \sum_{v=1}^{p_j} (\ln(l_{jv}))^2. \tag{6}$$

For testing $H_{0j} : f_{jv} = \text{constant}, \forall v \in \{1, \ldots, p_j\}$, we compare M6 with M5 via computing $\text{pBF}_j = \frac{1}{S}\sum_{s=1}^{S}(LP_{6j}^{(s)} - LP_{5j}^{(s)})/\ln(10)$, where all the quantities are defined similar to the previous subsection. We assign the mechanistic evidence $\mathcal{E}_{j2} = \text{pBF}_j$ for the gene of interest. As before, we use the following standard significance ranges for these quantities to categorize levels of evidence: $< 0.5$ (no evidence), $0.5 - 1$ (substantial), $1 - 2$ (strong), and $> 2$ (decisive).[4] This procedure is then performed independently for each of the $q$ genes, as described in Figure S1.

### S1.4. *Calibration function for Hierarchical Bayesian Variable Selection Model*

The calibration function $\mathcal{F}$ presented in Equation 4 in the main manuscript has to serve two primary purposes. First, it should be able to aggregate multi-dimensional prior evidence into a scalar prior probability, which means it needs to be a function on $\mathbb{R}^E \rightarrow [0, 1]$. Second, since the evidence quantities in our case belong to a continuous and nondecreasing spectrum of evidence strength, the function needs to preserve this unidirectional nature – increment in one source of evidence while keeping the others fixed should result in equal or higher calibrated prior probability. To serve these two purposes, we decompose the function as $\mathcal{F}(\mathcal{E}_j) = \mathcal{F}_1(\mathcal{F}_2(\mathcal{E}_j))$. $\mathcal{F}_2 : \mathbb{R}^E \rightarrow \mathbb{R}$ aggregates the multiple lines of evidence to a single scalar value $\bar{\mathcal{E}}_j$. We explore a linear map for this, as $\bar{\mathcal{E}}_j = \mathcal{F}_2(\mathcal{E}_j) = \sum_{e=1}^{E} \omega_{je}\mathcal{E}_{je}$. Here $\omega_{je}$s are convex weights specific to gene $j$, interpreted as quantifications of the importance of each source of evidence for that gene. Several choices of the $\omega_{je}$s are possible, as described below.

(1) **Average evidence:** $\omega_{je} = 1/E$ (takes a simple average of all available evidences).
(2) **Maximal evidence:** $\omega_{je} = I\big(\mathcal{E}_{je} = \max_{e' \in \{1,\ldots,E\}} \mathcal{E}_{je'}\big)$ (only takes into account the strongest evidence available from any source).
(3) **Precision-weighted evidence:** $\omega_{je} = \rho_{je}/\sum_{e=1}^{E} \rho_{je}$ (weights the evidences by some metric of reliability of the evidences, such as $\rho_{je} = \hat{\sigma}_{je}^{-2}$ where $\hat{\sigma}_{je}^2$ is the estimated noise variance for the

source model of $\mathcal{E}_{je}$).

$\mathcal{F}_1 : \mathbb{R} \to [0,1]$ maps the scalar evidence summary $\bar{\mathcal{E}}_j$ to the beta parameter. In our setting, this function is required to have the following features: for small positive or nonpositive $\bar{\mathcal{E}}_j$ (indicating small to no evidence for gene $j$) the beta parameter should be close to one, resulting in a prior distribution close to U(0, 1) for $\theta_j$; for larger values of $\bar{\mathcal{E}}_j$ the prior distribution should put increasing mass towards one. The rate of increase is guided by the cut-off ranges for the $\bar{\mathcal{E}}_j$s as described before.[4] To achieve this, we use $\mathcal{F}_1(\bar{\mathcal{E}}_j) = [[1 + \{\max(\bar{\mathcal{E}}_j, 10^{-6})/3\}^{-2.75}]^{-1} + 1]^4$. Using this function, the calibrated prior means of $\theta_j$ (representative values of $\bar{\mathcal{E}}_j$ in parentheses) are as follows: 0.502 (0.25), 0.543 (0.75), 0.726 (1.5), 0.962 (3). As illustrated in Supplementary Figure S2, the corresponding prior distributions of $\theta_j$ shift from an uniform prior to one concentrated close to one with increase in prior evidence strength.

### S1.5. *Variable Selection using False Discovery Rate Control*

Suppose the estimated posterior probabilities of inclusion are denoted by $\{\hat{\theta}_j, j \in \{1, \dots, q\}\}$. Then, we define p-value type quantities $p_j = 1 - \hat{\theta}_j, \forall j \in \{1, \dots, q\}$, and sort them in the increasing order of magnitude as $\{p_j^*, j \in \{1, \dots, q\}\}$, with the understanding that for each $j$, $p_j^* = p_{k_j}$ for some $k_j \in \{1, \dots, q\}$. Let the cumulative sums of these ordered quantities be denoted by $r_j = \sum_{l=1}^{j} p_j^*$ for each $j$. Let $j^* := \min\{j : r_j \geq \alpha\}$, where $\alpha$ is a pre-specified level for the false discovery rate control. Then we infer that the covariates with indices $k_1, \dots, k_{j^*}$ are selected in the outcome model.

### S1.6. *Cleaning and Filtering of Multiomic Cancer Cell Line and Patient Data*

**Filtering of Genes** Only the genes satisfying the following set of requirements in the cell lines data from CCLE are included in all analyses.

(1) Minimum sample size of 100 across breast, ovary, and uterus lineages.
(2) At least two matched upstream covariate (copy number or methylation) available in the dataset
(3) The coefficient of variation (CV, percentage scale) across the merged multi-lineage gene expression data is at least 25 (genes with too low CV have low variability in the samples of interest and are less informative for the second-stage cBVS model).

These cleaning steps result in a panel of 5,792 genes that pass all the tests. All these genes are included in the mechanistic and outcome model building procedures. Expression data for each gene is mean-centered before the analyses.

**Filtering of Drugs** Among the drugs available in the CCLE dataset, only those with at least 20 samples in all three lineages were included in the outcome model analyses, resulting in a total of 65 drugs/treatments. IC50 values (log-scale) are used as outcome variables in the drug response models, after mean centering for each drug × lineage combination.

### S1.7. *Uncalibrated Drug Response Models using Bayesian Model Averaging*

**Model specification** To assess the discovery performance of our calibrated Bayesian variable selection models in context of drug response, we construct uncalibrated drug response models using the Bayesian model averaging (BMA) procedure implemented in the *BMS R package*.[5] Recall the general form of our outcome models as described previously in Equation 3 in the main manuscript. Using the same notations therein, the uncalibrated Bayesian variable selection model explored by the BMA procedure can be written as the following.

$$Y_i = \sum_{j=1}^{q} \beta_j G_{ij} + \eta_i, i \in \{1, \dots, n\}. \tag{7}$$

Here $\eta_i$ are iid $N(0, \tau^2), \forall i \in \{1, ..., n\}$ as before. Now, $\boldsymbol{\beta} = (\beta_1, \dots, \beta_q)^T \sim \mathbf{N}_q(\mathbf{0}, g\tau^2(\boldsymbol{G}^T\boldsymbol{G})^{-1})$, which is Zellner's g-prior. For our implementations using BMA, we use the priors $\ln(\tau) \sim U(\mathbb{R})$ and $g = n$.

**Model Exploration** In context of this model, then, BMA estimates models for all possible combinations of the q genes of interest by constructing a weighted average on them. This means that the procedure estimates $\gamma \in \{1, \dots, 2^q\}$ many models (since each covariate may or may not be included in a model), where the model weights can be computed as the following using Bayes' theorem.

$$p(\text{Model}_\gamma | \boldsymbol{Y}, \boldsymbol{G}) = \frac{p(\boldsymbol{Y}|\text{Model}_\gamma, \boldsymbol{G}) p(\text{Model}_\gamma)}{p(\boldsymbol{Y}|\boldsymbol{G})} = \frac{p(\boldsymbol{Y}|\text{Model}_\gamma, \boldsymbol{G}) p(\text{Model}_\gamma)}{\sum_{s=1}^{2^q} p(\boldsymbol{Y}|\text{Model}_s, \boldsymbol{G}) p(\text{Model}_s)}. \tag{8}$$

Once we have computed these posterior model probabilities (PMPs), it is then possible to compute the model-weighted posterior distributions for any parameters or functions thereof. For example, if we are interested in the posterior of the coefficient $\beta_1$ corresponding to the first gene, we can compute the following.

$$p(\beta_1 | \boldsymbol{Y}, \boldsymbol{G}) = \sum_{s=1}^{2^q} p(\beta_1 | \text{Model}_s, \boldsymbol{Y}, \boldsymbol{G}) p(\text{Model}_s | \boldsymbol{Y}, \boldsymbol{G}). \tag{9}$$

Similar to the posterior distributions of the parameters, we can compute the posterior inclusion probabilities (PIPs) for a variable by summing up the PMPs for all models out of the $2^q$ where that variable was included. For example, the PIP for the first gene can be computed in the following way.

$$\text{PIP}_1 = \sum_{s=1}^{2^q} p(\text{Model}_s | \boldsymbol{Y}, \boldsymbol{G}) \boldsymbol{I}(\beta_1 \neq 0 | \text{Model}_s). \tag{10}$$

Similar calculations can then be extended to all the other genes. For the model priors $p(\text{Model}_\gamma)$, we use the default choice of setting $p(\text{Model}_\gamma) \propto 1$ i.e., uniform priors due to the lack of additional knowledge, as we did while computing the pBFs in our cBVS models. These PIPs can then be treated similar to $\hat{\theta}_j$s from the cBVS model and used in an FDR-controlled variable selection procedure as described in the previous subsection.

### S1.8. *Genes with Decisive Mechanistic Evidence from Cell Line Models but No Evidence from Any Patient Model*

ACKR1, ADAMTSL4, ADIRF, AGAP2-AS1, ANHX, AP4B1-AS1, ARHGEF25, ASCL5, BANF2, BARHL1, BHLHE23, BSPH1, C5orf47, C5orf66-AS1, C9orf92, CA4, CADM3-AS1, CAV3, CD248, COL6A5, CRACR2B, CRYAA, CSF3, CYGB, CYSRT1, DDX11L1, DDX4, DDX53, DLG1-AS1, DNAH17-AS1, DRD5, EGFR-AS1, ELDR, ELFN1-AS1, EMC10, ENKD1, ERVFRD-1, EVA1B, FABP5P3, FABP9, FAM210B, FAM221A, FAM72C, FAM99B, FKBP11, FUOM, GACAT2, GIMAP5, GTSF1, HAPLN4, HCAR1, HENMT1, HIPK1-AS1, HMX1, HOXA4, HOXB-AS1, HOXC-AS2, HSD17B1, HTATSF1P2, HTR5A, HTR5A-AS1, IL3, ILDR2, INSM2, ITGA5, KCNA10, KCTD21-AS1, KDF1, KRT27, LCE1B, LCE2D, LHX5-AS1, LINC00200, LINC00226, LINC00523, LINC00526, LINC00528, LINC00624, LINC00662, LINC00665, LINC00667, LINC00682, LINC00884, LINC00899, LINC01019, LINC01044, LINC01082, LINC01090, LINC01097, LINC01101, LINC01182, LINC01230, LINC01276, LINC01346, LINC01354, LINC01426, LINC01427, LRRC38, MBD3L3, MCHR2-AS1, MCTS2P, MEIOB, MIA-RAB4B, MICU3, MIEN1, MIR181D, MIRLET7BHG, MISP, MOS, MRGPRG, MRGPRG-AS1, MROH5, MSANTD4, MSC, MUC17, MYL9, NANOGNB, NAPRT, NDUFC2-KCTD14, NEUROD1, NEXN, NEXN-AS1, NKX2-6, NMRK2, NNT-AS1, NOL4L, NWD2, OR10V1, OR10X1, OR1Q1, OR2T11, OR2T12, OR5D18, OR5P3, OR6K6, PCAT7, PCDHA4, PCDHGA11, PCDHGB5, PIEZO2, PLBD1-AS1, POU3F4, PRND, PRR34-AS1, PRRT3-AS1, PRSS56, PWRN2, PXDC1, RAD21-AS1, RARA-AS1, RBBP8NL, REC114, RHOV, RIPPLY3, RNF223, RNF225, SAGE1, SCT, SIX6, SLC25A21-AS1, SLC35G2, SLC36A2, SLC9A7P1, SMCO4, SMIM22, SNORA70B, SNORA81, SOWAHA, SPATA8-AS1, SPATC1L, ST8SIA6-AS1, SWT1, TAC4, TBC1D3P1-DHX40P1, TCF21, TDRP, TENM4, TEX11, THEMIS2, THOC7-AS1, TMEM247, TOR4A, TRIQK, TRPM2-AS, TSPY26P, TSPYL6, TTC39A-AS1, TUBA4B, TYRO3P, UBE2DNL, VGLL2, VIM, VPS9D1-AS1, VWC2L, WWC2-AS2, ZC2HC1A, ZFP36, ZKSCAN7, ZNF300P1, ZNF385D-AS1, ZNF486, ZNF529-AS1, ZNF542P, ZNF571-AS1, ZNF728, ZNF790-AS1, ZNRF4, ZSCAN30.

### S2. Supplementary Tables

| Cell Lines Data (CCLE and GDSC) | | |
|---|---|---|
| **Data** | **Platform** | **Download Link** |
| Gene expression | RNAseq TPM RSEM<br>(log2 transformed using a pseudo-count of 1) | Link |
| Copy Number | Gene-level (log2 transformed with a pseudo-count of 1).<br>Inferred from WGS, WES or SNP array depending on availability.<br>Calculated by mapping genes onto segment level calls and computing a weighted average. | Link |
| Methylation | Reduced representation bisulfite sequencing (promoter CpG clusters). | Link |
| Drug Response | Multiple dose-response parameters available.<br>IC50s used. | Link |
| Metadata | — | Link |
| **Patient Data (TCGA)** | | |
| **Data** | **Platform** | **Download Link** |
| Gene Expression | Illumina HiSeq 2000 (log2 transformed RSEM normalized count). | BRCA<br>UCS<br>OV |
| Copy Number | Gene-level copy number variation (CNV) estimated using GISTIC2. | BRCA<br>UCS<br>OV |
| Methylation | Illumina Infinium HumanMethylation450 platform (BRCA, UCS)<br>Illumina Infinium HumanMethylation27 platform (OV) | BRCA<br>UCS<br>OV |

**Supplementary Table S1: Summary information on sources and platforms for multiomic cancer cell line and patient data.** Only 10 samples are available for the Methylation450 platform for OV, which is why Methylation27 was used instead.

| Patient Model | | | Genes |
|---|---|---|---|
| **BRCA** | **UCS** | **OV** | |
| No Evidence | No Evidence | No Evidence | See Supplementary Notes Section S1.8 |
| Substantial | No Evidence | No Evidence | ADAMTS4, PCDHGA3, RTP3, SMC1B |
| No Evidence | Strong | No Evidence | AGBL1, CNPY1, PLA2G12B |
| No Evidence | No Evidence | Strong | APOC2, C16orf86, UPK3B, ZNF626 |
| Substantial | Substantial | No Evidence | AQP12B |
| No Evidence | Substantial | No Evidence | CASR, MORC1 |
| Strong | No Evidence | No Evidence | CCT8L2, HES5, MAB21L1, MFAP2 |
| No Evidence | No Evidence | Substantial | ELF3, PCDHGA8, PLBD1 |
| Strong | No Evidence | Strong | IER3 |
| Strong | No Evidence | Substantial | RIN1 |
| No Evidence | Strong | Strong | ZNF880 |

**Supplementary Table S2: Mechanistic evidence summary for genes with decisive evidence only from the cell lines multi-lineage model.**

| Gene | PIP | CL | BRCA | OV | UCS |
|---|---|---|---|---|---|
| LIPH | 0.9995 | No Evidence | Decisive | Decisive | No Evidence |
| ZNF728 | 0.9992 | Decisive | No Evidence | No Evidence | No Evidence |
| TNFRSF25 | 0.9992 | No Evidence | Decisive | Decisive | No Evidence |
| BTG3 | 0.9986 | Decisive | Decisive | Decisive | Strong |
| ANXA9 | 0.9984 | Decisive | Decisive | Decisive | Strong |
| ZIC4 | 0.9984 | No Evidence | Decisive | No Evidence | No Evidence |
| CD83 | 0.9983 | No Evidence | Decisive | No Evidence | No Evidence |
| BLMH | 0.9983 | No Evidence | Decisive | Decisive | No Evidence |
| SLIT3 | 0.9982 | No Evidence | Decisive | No Evidence | Strong |
| ITPR1 | 0.9982 | No Evidence | Decisive | No Evidence | Substantial |
| CTSC | 0.9982 | Strong | Decisive | Strong | Decisive |
| KIAA1614 | 0.9981 | No Evidence | Decisive | No Evidence | Strong |
| GRK5 | 0.9979 | No Evidence | Decisive | No Evidence | Strong |
| ADAM19 | 0.9978 | Decisive | Decisive | No Evidence | Strong |
| ZBTB42 | 0.9977 | No Evidence | Decisive | Decisive | Strong |
| HDAC11 | 0.9977 | No Evidence | Decisive | Decisive | Substantial |
| MICAL1 | 0.9975 | No Evidence | Decisive | No Evidence | Strong |
| CNFN | 0.9975 | No Evidence | Decisive | Decisive | Strong |
| S1PR2 | 0.9974 | No Evidence | Decisive | Decisive | No Evidence |
| FSCN1 | 0.9974 | No Evidence | Decisive | Strong | Decisive |
| RHOB | 0.9974 | No Evidence | Decisive | Substantial | Substantial |
| MFGE8 | 0.9973 | No Evidence | Decisive | Substantial | Substantial |
| PRR15 | 0.9973 | Decisive | Decisive | No Evidence | No Evidence |
| TAF4B | 0.9972 | No Evidence | Decisive | Decisive | No Evidence |
| PKNOX2 | 0.9971 | No Evidence | Decisive | No Evidence | Decisive |
| LRRCC1 | 0.9971 | No Evidence | Decisive | Decisive | No Evidence |
| AEBP1 | 0.9971 | Decisive | Decisive | No Evidence | No Evidence |
| SCML2 | 0.997 | No Evidence | Decisive | No Evidence | Strong |
| LZTS1 | 0.997 | No Evidence | Decisive | No Evidence | Substantial |
| FAT2 | 0.997 | No Evidence | Decisive | No Evidence | No Evidence |
| PTK6 | 0.997 | Decisive | Decisive | Decisive | Strong |
| HAL | 0.9968 | No Evidence | Decisive | Decisive | No Evidence |
| ACVRL1 | 0.9968 | No Evidence | Decisive | No Evidence | Strong |
| TMEM51 | 0.9968 | No Evidence | Decisive | Decisive | Substantial |
| SLC6A5 | 0.9967 | Decisive | No Evidence | Decisive | No Evidence |
| ZNF670 | 0.9967 | No Evidence | Decisive | Decisive | No Evidence |
| NEBL | 0.9966 | No Evidence | Decisive | Strong | Substantial |
| SHISA2 | 0.9966 | No Evidence | Decisive | No Evidence | Decisive |
| CNGA3 | 0.9966 | No Evidence | Decisive | No Evidence | Substantial |
| RAB39B | 0.9966 | No Evidence | Decisive | No Evidence | No Evidence |

| Gene | PIP | CL | BRCA | OV | UCS |
|---|---|---|---|---|---|
| WBP2NL | 0.9966 | No Evidence | Decisive | No Evidence | Strong |
| BAHCC1 | 0.9965 | No Evidence | Decisive | No Evidence | Decisive |
| CXXC5 | 0.9965 | No Evidence | Decisive | Decisive | Decisive |
| PRSS41 | 0.9965 | Strong | Decisive | No Evidence | Strong |
| SUSD3 | 0.9965 | Decisive | Decisive | Decisive | Substantial |
| GATA3 | 0.9965 | Decisive | Decisive | No Evidence | Decisive |
| SDR16C5 | 0.9964 | No Evidence | Decisive | No Evidence | Strong |
| METRN | 0.9964 | Decisive | Decisive | No Evidence | No Evidence |
| ST8SIA6 | 0.9964 | No Evidence | Decisive | No Evidence | No Evidence |
| MYRIP | 0.9963 | No Evidence | Decisive | Substantial | Strong |
| BTN1A1 | 0.9963 | No Evidence | Decisive | No Evidence | No Evidence |
| FUT11 | 0.9963 | No Evidence | Decisive | Decisive | No Evidence |
| SLC23A1 | 0.9963 | No Evidence | Decisive | Substantial | Substantial |
| ZNF283 | 0.9962 | No Evidence | Decisive | Decisive | Substantial |
| TMEM65 | 0.9962 | Decisive | Decisive | Decisive | Strong |
| KIF21B | 0.9962 | No Evidence | Decisive | No Evidence | Substantial |
| DNAJC5B | 0.9962 | No Evidence | Decisive | No Evidence | No Evidence |
| APBB2 | 0.9962 | No Evidence | Decisive | Strong | No Evidence |
| VAMP1 | 0.9962 | No Evidence | Decisive | Decisive | No Evidence |
| LINC00226 | 0.9962 | Decisive | No Evidence | No Evidence | No Evidence |
| LRP12 | 0.9962 | No Evidence | Decisive | Decisive | Substantial |
| MIPOL1 | 0.9962 | Decisive | Decisive | Strong | Substantial |
| GPC2 | 0.9962 | No Evidence | Decisive | No Evidence | No Evidence |
| PRKCA | 0.9961 | No Evidence | Decisive | Strong | Strong |
| ISYNA1 | 0.9961 | Decisive | Decisive | Decisive | Strong |
| RGS20 | 0.9961 | No Evidence | Decisive | Strong | Substantial |
| ANGPT1 | 0.9961 | No Evidence | Decisive | No Evidence | No Evidence |

**Supplementary Table S3: Summary for genes selected in the docetaxel response model for breast cell lines.** PIP denotes the posterior inclusion probability in the calibrated Bayesian variable selection model. The last four columns indicate the level of mechanistic evidence determined by the pBFs from the corresponding models for that gene.

| Gene | PIP | CL | BRCA | OV | UCS |
|---|---|---|---|---|---|
| CDCA7 | 0.9992 | Strong | Decisive | No Evidence | No Evidence |
| SLC24A3 | 0.9986 | No Evidence | Decisive | No Evidence | No Evidence |
| SLC27A5 | 0.9986 | No Evidence | Decisive | Decisive | Strong |
| PXDC1 | 0.9986 | Decisive | No Evidence | No Evidence | No Evidence |
| PLVAP | 0.9985 | No Evidence | Decisive | No Evidence | No Evidence |
| MUC4 | 0.9985 | No Evidence | Decisive | Strong | No Evidence |
| TNFAIP2 | 0.9983 | Decisive | Decisive | Decisive | Substantial |
| CYBA | 0.9982 | Decisive | Decisive | Decisive | Strong |
| TBL1X | 0.9982 | No Evidence | Decisive | No Evidence | Decisive |
| TRIM21 | 0.9979 | No Evidence | Decisive | Decisive | Substantial |
| EPPK1 | 0.9979 | Decisive | Decisive | Decisive | Strong |
| CROT | 0.9978 | Decisive | No Evidence | Decisive | Substantial |
| PTH2R | 0.9978 | Decisive | Decisive | No Evidence | Decisive |
| BANK1 | 0.9978 | No Evidence | Decisive | Decisive | Strong |
| PRR18 | 0.9978 | No Evidence | Decisive | No Evidence | Strong |
| KLF2 | 0.9977 | No Evidence | Decisive | No Evidence | No Evidence |
| MGST2 | 0.9977 | Decisive | Decisive | Decisive | Strong |
| CADM1 | 0.9977 | No Evidence | Decisive | Decisive | Decisive |
| ZNF652 | 0.9977 | Decisive | Decisive | Decisive | Strong |
| ZNF506 | 0.9977 | Decisive | Decisive | Decisive | No Evidence |
| PCDHGB4 | 0.9976 | No Evidence | No Evidence | Decisive | No Evidence |
| ESPN | 0.9976 | No Evidence | Decisive | Decisive | Decisive |
| ZNF572 | 0.9976 | Decisive | Decisive | Decisive | Decisive |
| CST6 | 0.9976 | Decisive | Decisive | No Evidence | No Evidence |
| LTB | 0.9976 | No Evidence | Decisive | No Evidence | No Evidence |
| ITGA3 | 0.9976 | No Evidence | Decisive | Decisive | Strong |
| PODN | 0.9975 | No Evidence | Decisive | No Evidence | No Evidence |
| RGS2 | 0.9975 | No Evidence | Decisive | No Evidence | No Evidence |
| SRR | 0.9975 | No Evidence | Decisive | Decisive | No Evidence |
| FZD9 | 0.9975 | No Evidence | Decisive | Decisive | No Evidence |
| ATP2B2 | 0.9975 | No Evidence | Decisive | Decisive | Decisive |
| MICAL2 | 0.9974 | No Evidence | Decisive | Decisive | No Evidence |
| TNFAIP3 | 0.9974 | No Evidence | Decisive | No Evidence | Strong |
| TRPV2 | 0.9973 | Decisive | Decisive | No Evidence | No Evidence |
| PHLDB2 | 0.9973 | No Evidence | Decisive | No Evidence | Strong |
| ASS1 | 0.9973 | Decisive | Decisive | Decisive | Decisive |
| HOXB7 | 0.9972 | No Evidence | Decisive | No Evidence | Substantial |
| C5orf38 | 0.9972 | Decisive | Decisive | No Evidence | No Evidence |
| EPHA2 | 0.9972 | Decisive | Decisive | Decisive | No Evidence |
| LY6K | 0.9972 | Decisive | Decisive | Decisive | Decisive |

| Gene | PIP | CL | BRCA | OV | UCS |
|---|---|---|---|---|---|
| BLMH | 0.9971 | No Evidence | Decisive | Decisive | No Evidence |
| EPHB1 | 0.9971 | No Evidence | Decisive | No Evidence | Decisive |
| BCL2L11 | 0.997 | No Evidence | Decisive | Decisive | No Evidence |
| SMIM22 | 0.997 | Decisive | No Evidence | No Evidence | No Evidence |
| F11R | 0.997 | Strong | Decisive | Decisive | No Evidence |
| CMTM3 | 0.997 | Decisive | Decisive | Decisive | No Evidence |
| IL15 | 0.997 | No Evidence | Decisive | Decisive | No Evidence |
| SLC16A7 | 0.997 | No Evidence | Decisive | Decisive | No Evidence |
| DLX1 | 0.997 | No Evidence | Decisive | Decisive | No Evidence |
| LHFPL2 | 0.997 | No Evidence | Decisive | Substantial | Substantial |
| PAK6 | 0.997 | No Evidence | Decisive | Decisive | Strong |
| PTPRZ1 | 0.9969 | No Evidence | Decisive | No Evidence | Decisive |
| SEMA6B | 0.9969 | Strong | Decisive | Decisive | Decisive |
| PPM1M | 0.9969 | No Evidence | Decisive | Substantial | No Evidence |
| ZNF425 | 0.9969 | Strong | Decisive | Decisive | Substantial |
| TET1 | 0.9969 | No Evidence | Decisive | No Evidence | No Evidence |
| EPB41L4A | 0.9969 | No Evidence | Decisive | Decisive | Strong |
| KRT8 | 0.9968 | No Evidence | Decisive | Decisive | Decisive |
| DENND1B | 0.9967 | No Evidence | Decisive | Decisive | Substantial |
| OR2T11 | 0.9967 | Decisive | No Evidence | No Evidence | No Evidence |
| EVPLL | 0.9967 | No Evidence | Decisive | No Evidence | No Evidence |
| SLCO6A1 | 0.9966 | No Evidence | Decisive | No Evidence | No Evidence |
| IFI27 | 0.9966 | Decisive | Decisive | No Evidence | Decisive |
| GDPD3 | 0.9966 | No Evidence | Decisive | Strong | No Evidence |
| AGBL1 | 0.9966 | Decisive | No Evidence | No Evidence | Strong |
| HOXC4 | 0.9966 | No Evidence | Decisive | Decisive | Decisive |
| SDR42E1 | 0.9966 | Decisive | Decisive | No Evidence | Decisive |
| SUSD2 | 0.9966 | No Evidence | Decisive | Decisive | No Evidence |
| NUPR1 | 0.9966 | Decisive | Decisive | No Evidence | Strong |
| MCTS2P | 0.9966 | Decisive | No Evidence | No Evidence | No Evidence |
| B4GALT6 | 0.9965 | No Evidence | Decisive | Decisive | No Evidence |
| ST14 | 0.9965 | Decisive | Decisive | Decisive | Decisive |
| NPEPL1 | 0.9965 | No Evidence | Decisive | Strong | No Evidence |
| TMC4 | 0.9965 | No Evidence | Decisive | Decisive | No Evidence |

**Supplementary Table S4: Summary for genes selected in the cisplatin response model for ovarian cell lines.** PIP denotes the posterior inclusion probability in the calibrated Bayesian variable selection model. The last four columns indicate the level of mechanistic evidence determined by the pBFs from the corresponding models for that gene.

### S3. Supplementary Figures

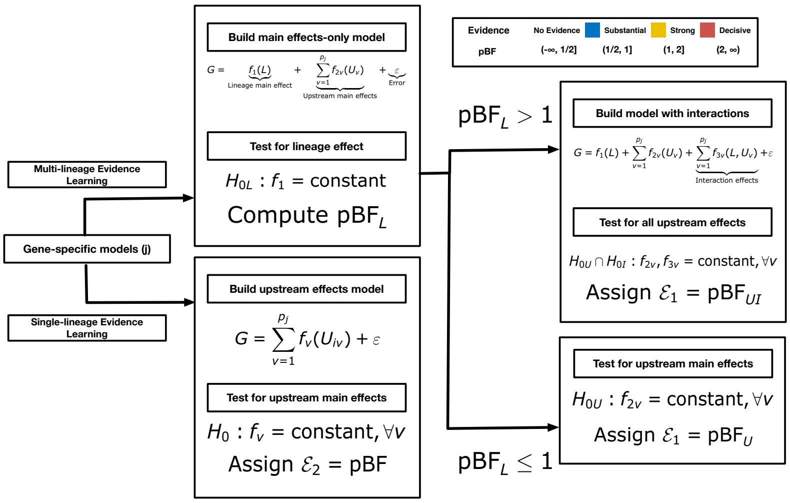


**Supplementary Figure S1: The sequential evidence detection procedure for identifying driver genes within the mechanistic layer of the *BaySyn* framework.** All pBFs are assumed to be at the $\log_{10}(\bullet)$-scale.

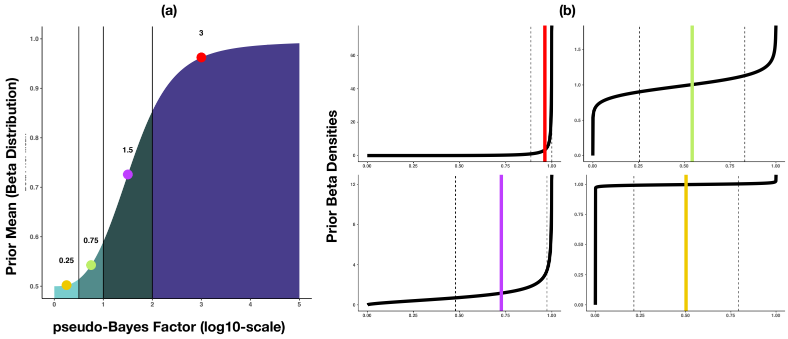


**Supplementary Figure S2: Calibration function for the calibrated Bayesian variable selection models.** The left panel plots the calibrated prior Beta means against the pseudo-Bayes factors. The right panel presents the densities (along with markers for means (solid vertical lines) and standard deviations (SD, broken vertical lines indicate the ±1 SD ranges around the means)) for four representative values from the four ranges of interest in the x-axis of the left panel.

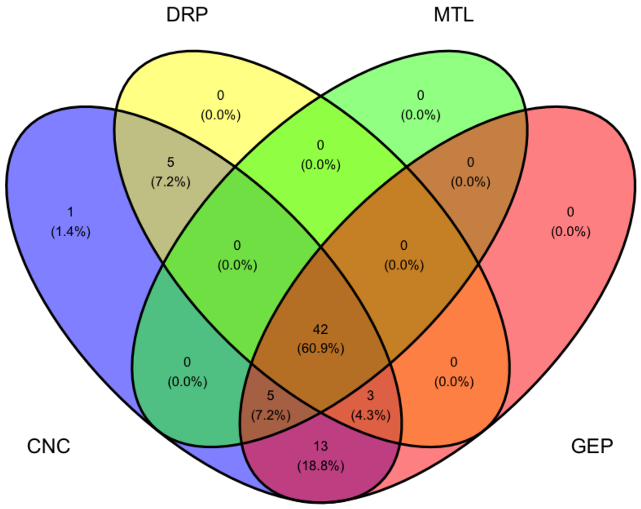


**Supplementary Figure S3: Sample size summaries across the different platforms for breast cell lines from CCLE.** The acronyms for the different data platforms are as the following - CNC: copy number, GEP: gene expression, MTL: DNA methylation, DRP: drug response. Specific information on each platform are available in Supplementary Table S1.

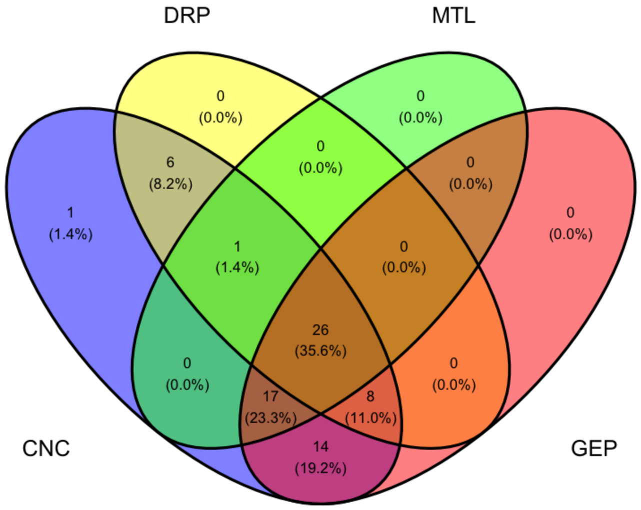


**Supplementary Figure S4: Sample size summaries across the different platforms for ovary cell lines from CCLE.** The acronyms for the different data platforms are as the following - CNC: copy number, GEP: gene expression, MTL: DNA methylation, DRP: drug response. Specific information on each platform are available in Supplementary Table S1.

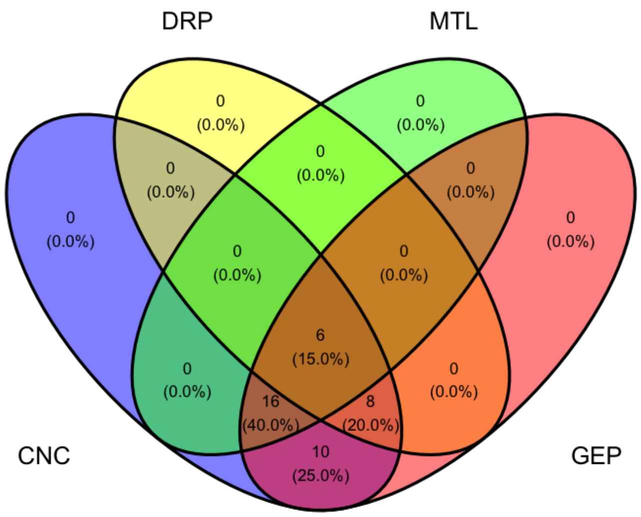


**Supplementary Figure S5: Sample size summaries across the different platforms for uterus cell lines from CCLE.** The acronyms for the different data platforms are as the following - CNC: copy number, GEP: gene expression, MTL: DNA methylation, DRP: drug response. Specific information on each platform are available in Supplementary Table S1.

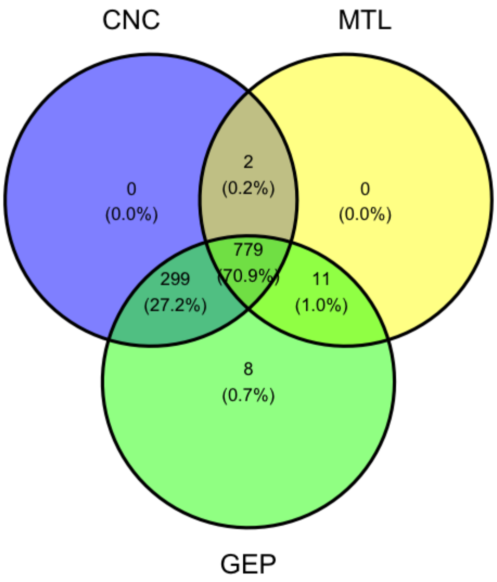


**Supplementary Figure S6: Sample size summaries across the different platforms for BRCA patients from TCGA.** The acronyms for the different data platforms are as the following - CNC: copy number, GEP: gene expression, MTL: DNA methylation. Specific information on each platform are available in Supplementary Table S1.

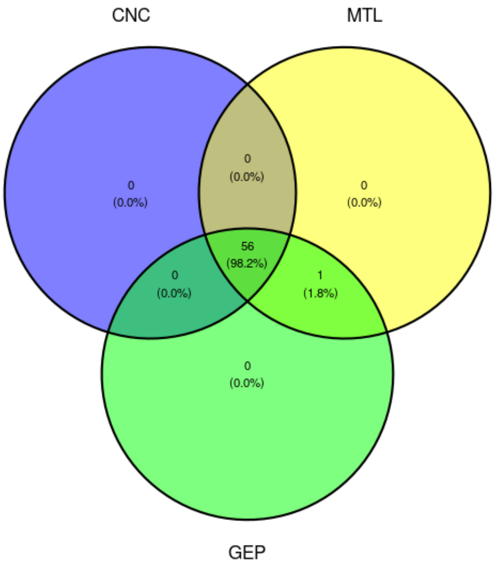


**Supplementary Figure S7: Sample size summaries across the different platforms for UCS patients from TCGA.** The acronyms for the different data platforms are as the following - CNC: copy number, GEP: gene expression, MTL: DNA methylation. Specific information on each platform are available in Supplementary Table S1.

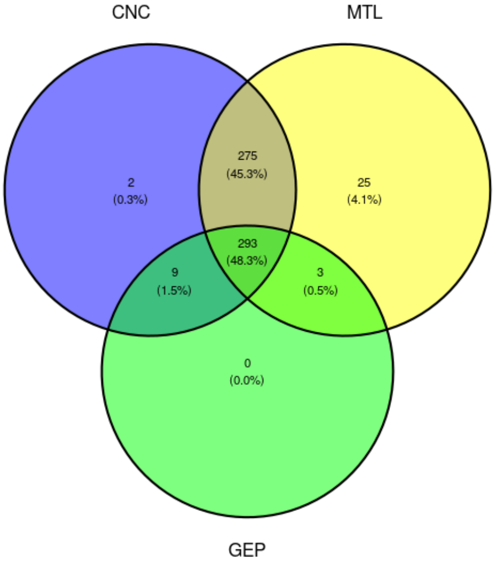


**Supplementary Figure S8: Sample size summaries across the different platforms for OV patients from TCGA.** The acronyms for the different data platforms are as the following - CNC: copy number, GEP: gene expression, MTL: DNA methylation. Specific information on each platform are available in Supplementary Table S1.

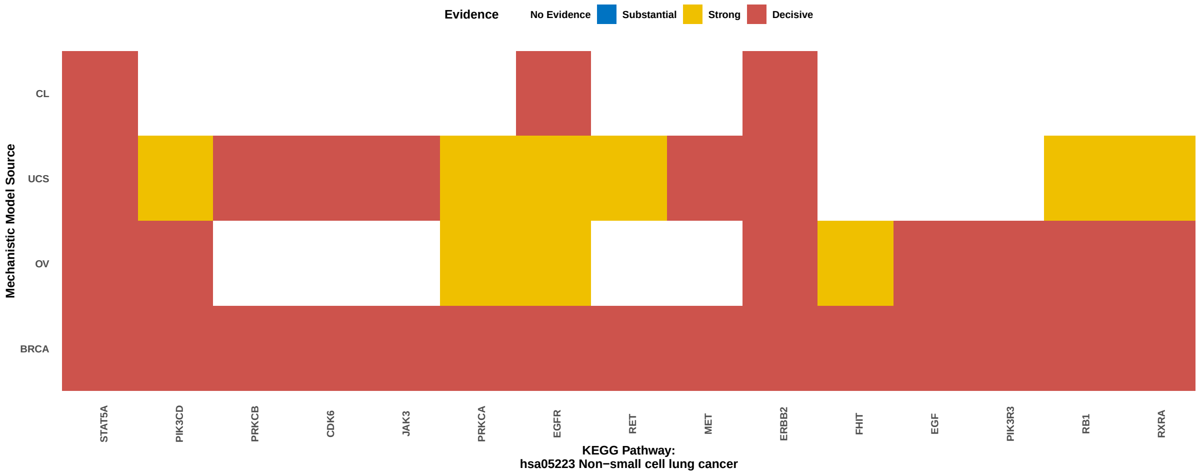


**Supplementary Figure S9: Heatmap summarizing levels of mechanistic evidence for the genes in KEGG non-small cell lung cancer gene set.** Genes in the rows are ordered based on clusters resulting from the evidence statistics.

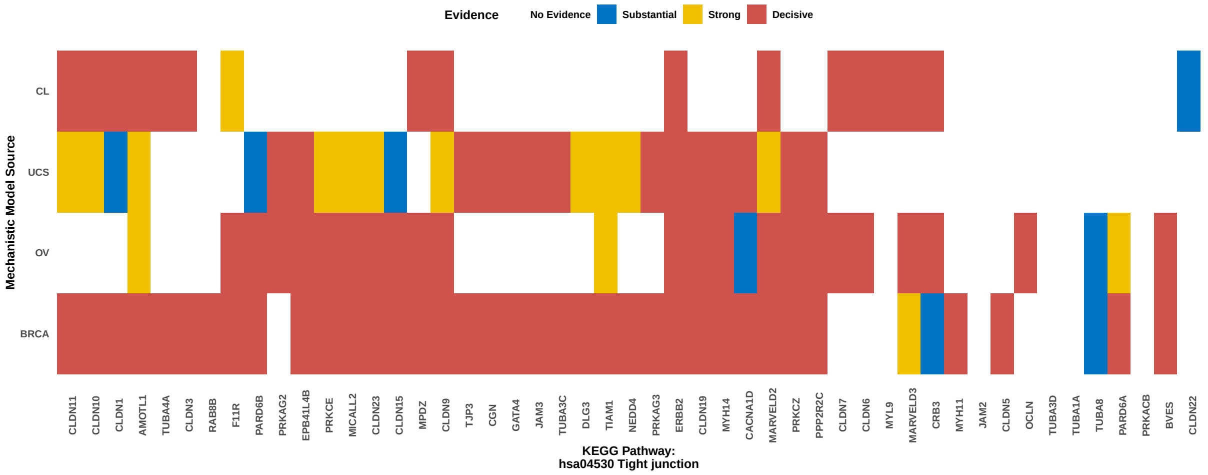


**Supplementary Figure S10: Heatmap summarizing levels of mechanistic evidence for the genes in KEGG tight junction gene set.** Genes in the rows are ordered based on clusters resulting from the evidence statistics.

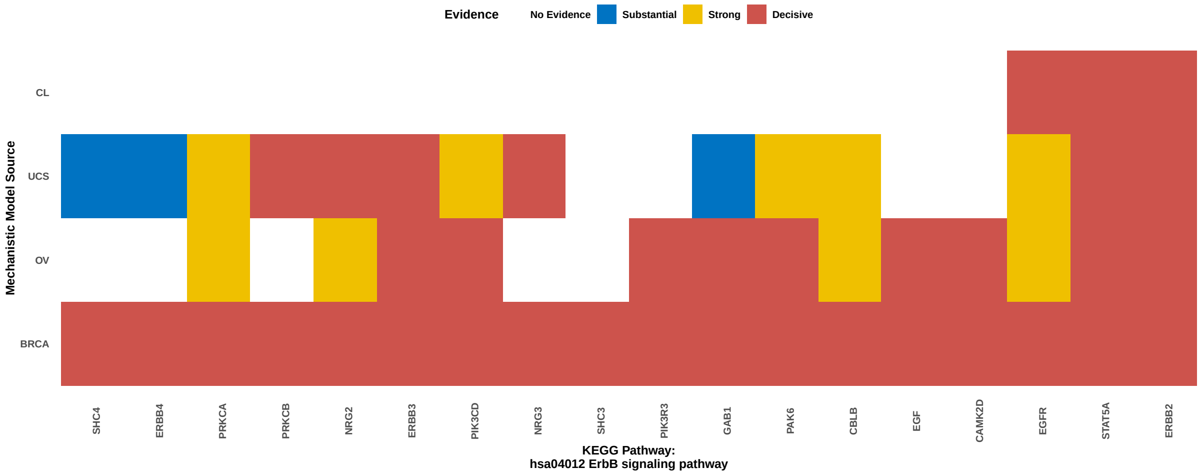


**Supplementary Figure S11: Heatmap summarizing levels of mechanistic evidence for the genes in KEGG ERBB signaling gene set.** Genes in the rows are ordered based on clusters resulting from the evidence statistics.

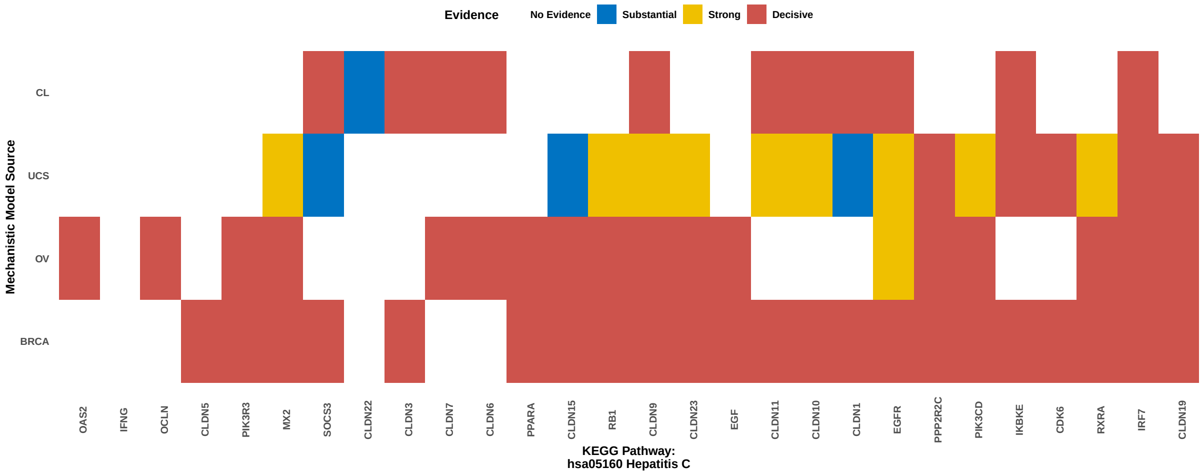


**Supplementary Figure S12: Heatmap summarizing levels of mechanistic evidence for the genes in KEGG hepatitis C gene set.** Genes in the rows are ordered based on clusters resulting from the evidence statistics.

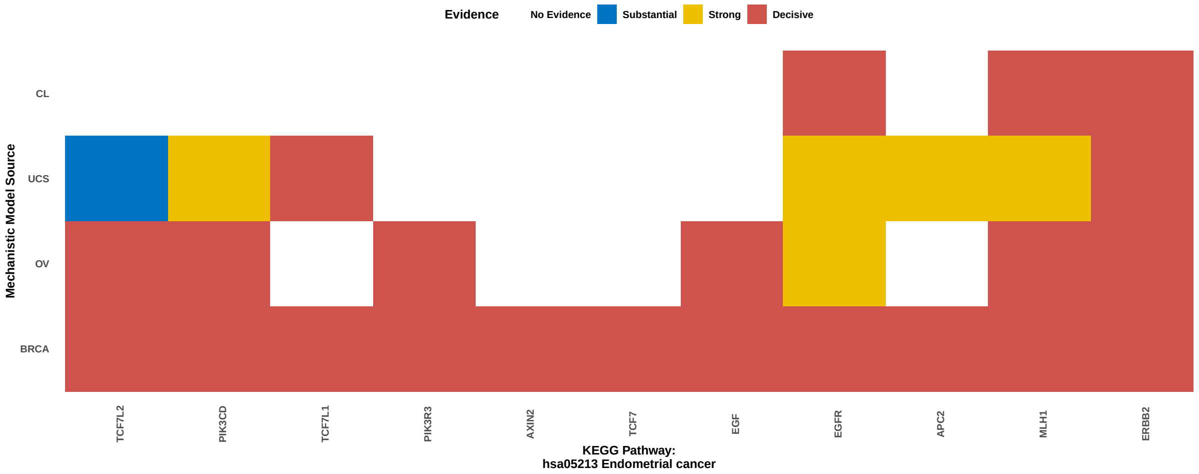


**Supplementary Figure S13: Heatmap summarizing levels of mechanistic evidence for the genes in KEGG endometrial cancer gene set.** Genes in the rows are ordered based on clusters resulting from the evidence statistics.

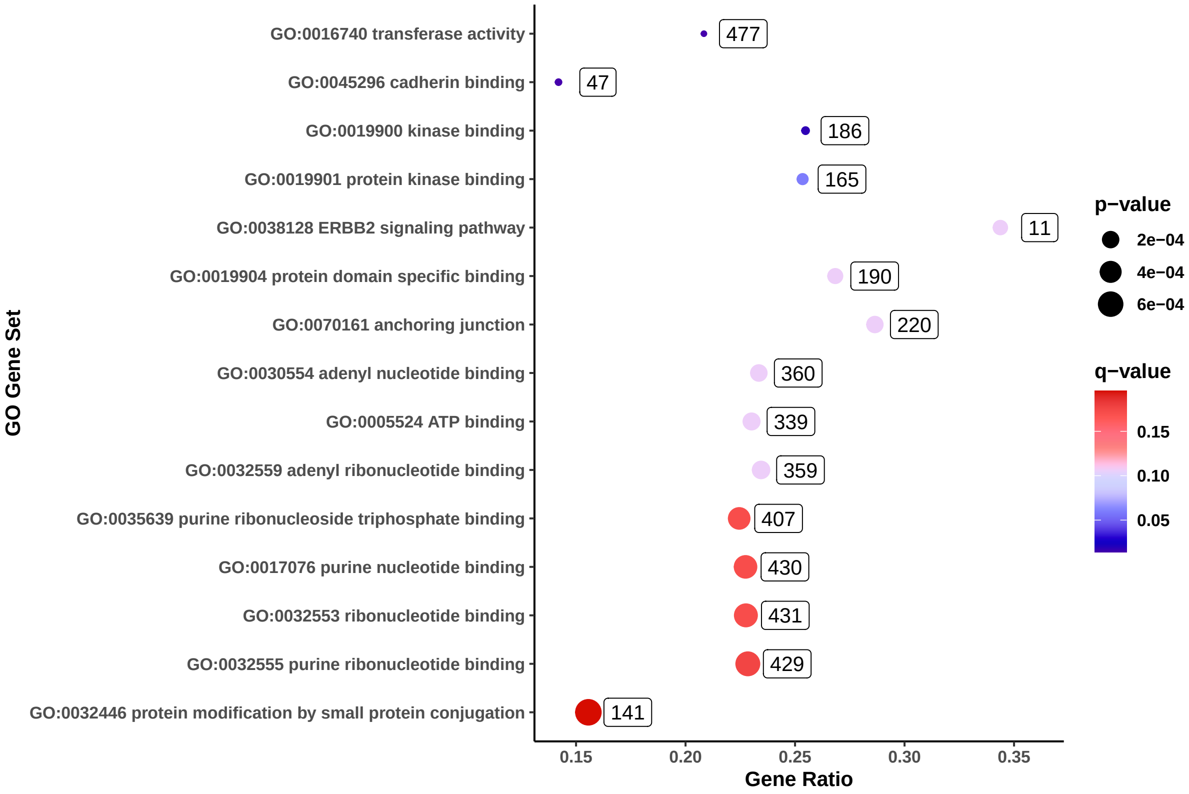


**Supplementary Figure S14: Dotplot summarizing significance levels for GO gene sets.** Only the sets with a q-value of at most 0.2 are included. The gene sets are ordered from top to bottom in decreasing order of q-values. The labels beside the dots indicate set sizes in our analyses.

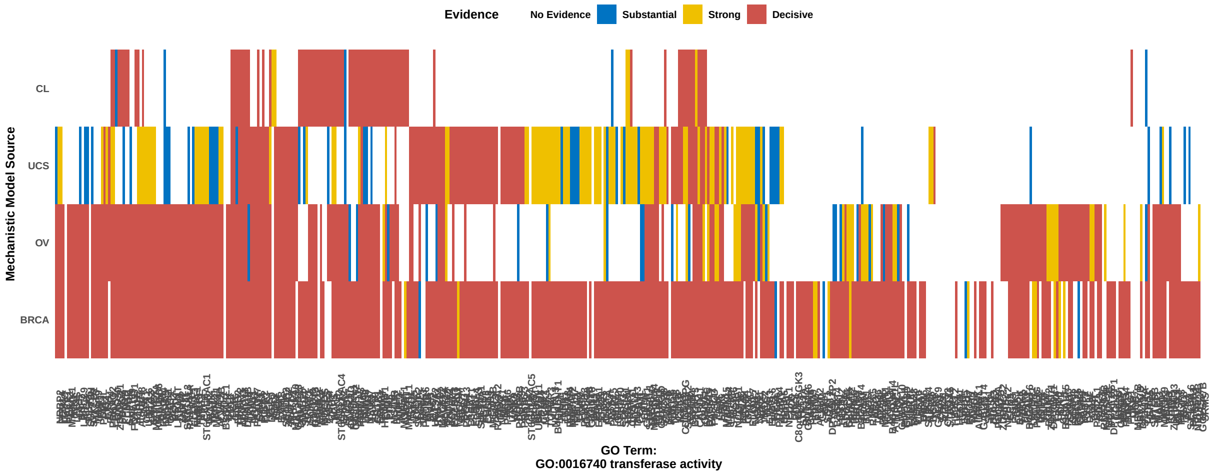


**Supplementary Figure S15: Heatmap summarizing levels of mechanistic evidence for the genes in GO transferase activity gene set.** Genes in the rows are ordered based on clusters resulting from the evidence statistics.

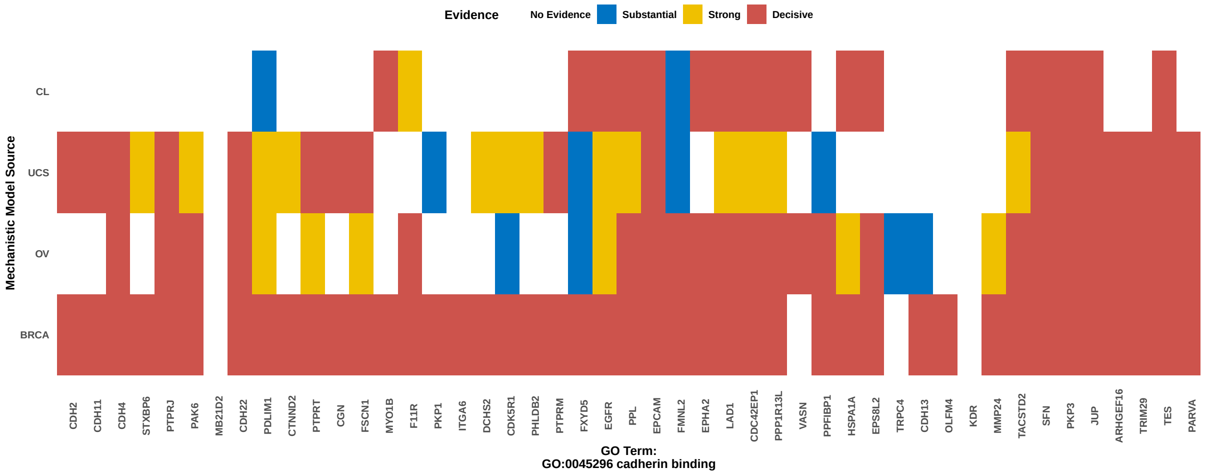


**Supplementary Figure S16: Heatmap summarizing levels of mechanistic evidence for the genes in GO cadherin binding gene set.** Genes in the rows are ordered based on clusters resulting from the evidence statistics.

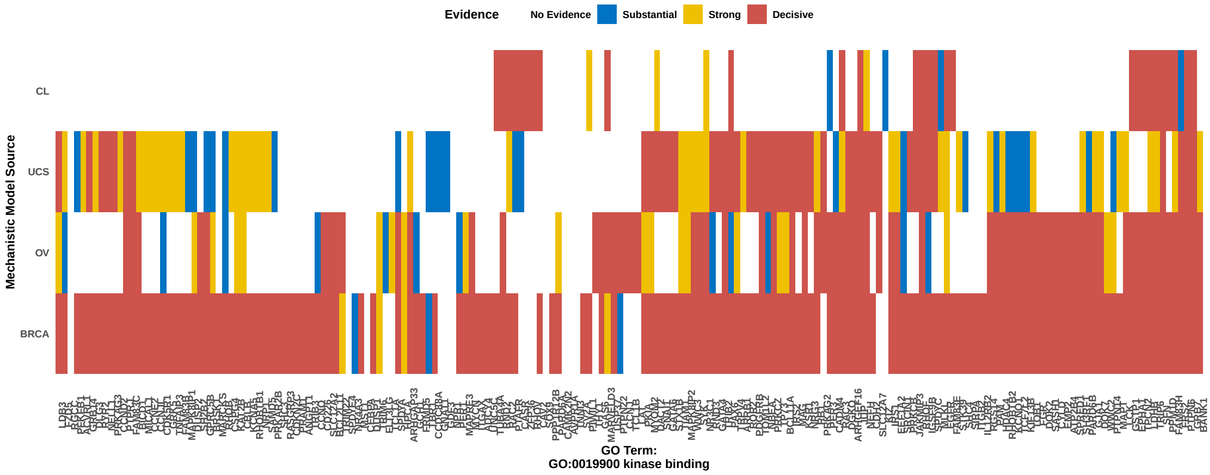


**Supplementary Figure S17: Heatmap summarizing levels of mechanistic evidence for the genes in GO kinase binding gene set.** Genes in the rows are ordered based on clusters resulting from the evidence statistics.

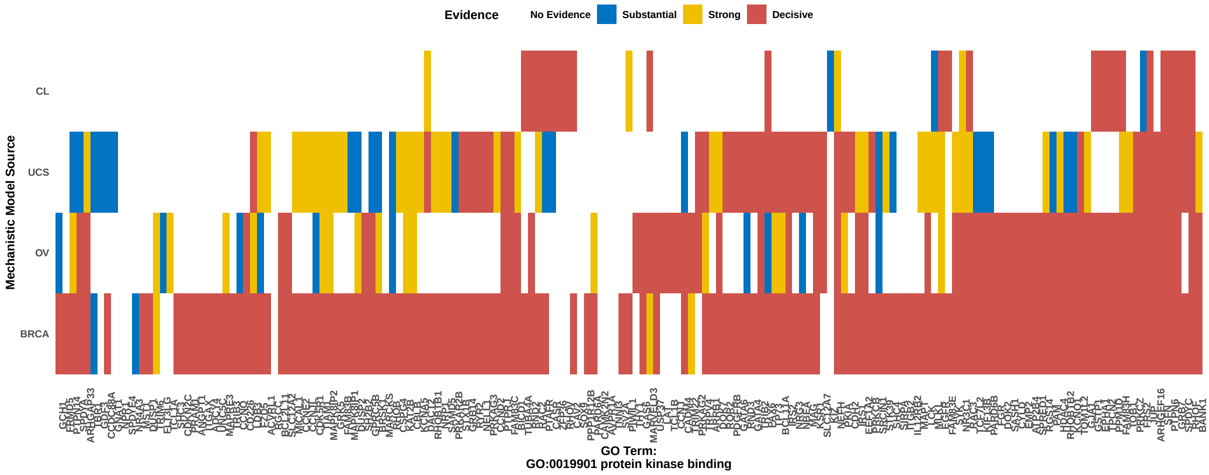


**Supplementary Figure S18: Heatmap summarizing levels of mechanistic evidence for the genes in GO protein kinase binding gene set.** Genes in the rows are ordered based on clusters resulting from the evidence statistics.

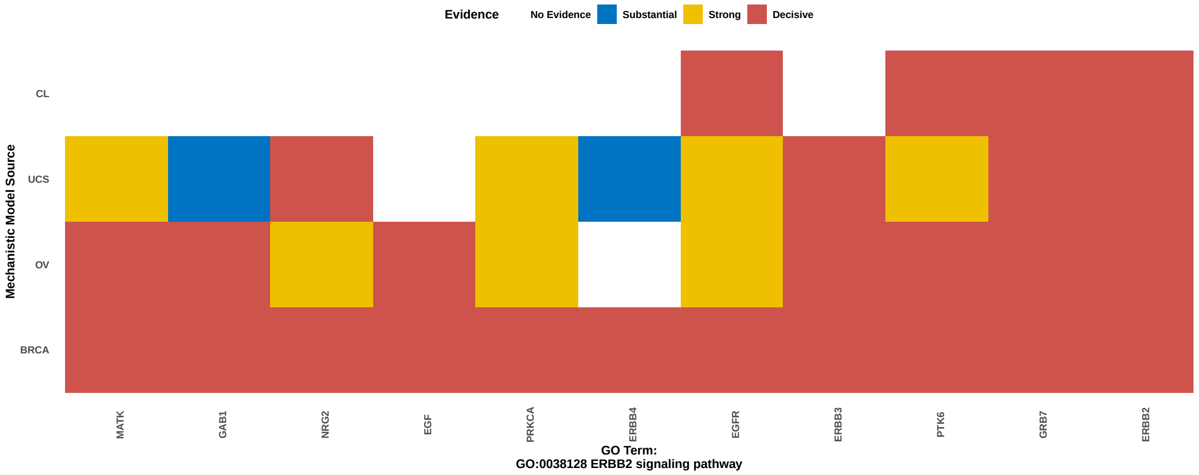


**Supplementary Figure S19: Heatmap summarizing levels of mechanistic evidence for the genes in GO ERBB2 signaling pathway gene set.** Genes in the rows are ordered based on clusters resulting from the evidence statistics.

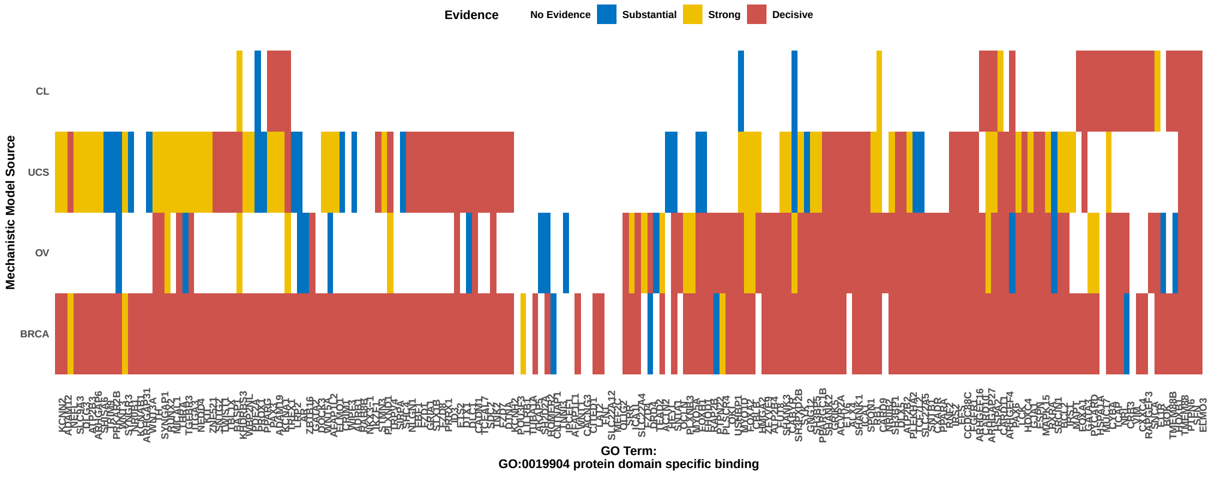


**Supplementary Figure S20: Heatmap summarizing levels of mechanistic evidence for the genes in GO protein domain specific binding gene set.** Genes in the rows are ordered based on clusters resulting from the evidence statistics.

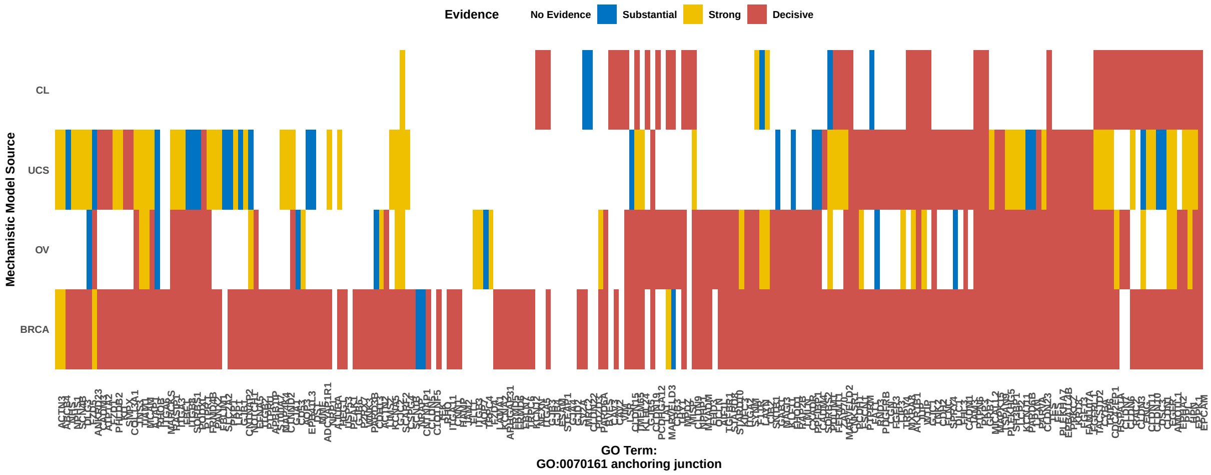


**Supplementary Figure S21: Heatmap summarizing levels of mechanistic evidence for the genes in GO anchoring junction gene set.** Genes in the rows are ordered based on clusters resulting from the evidence statistics.

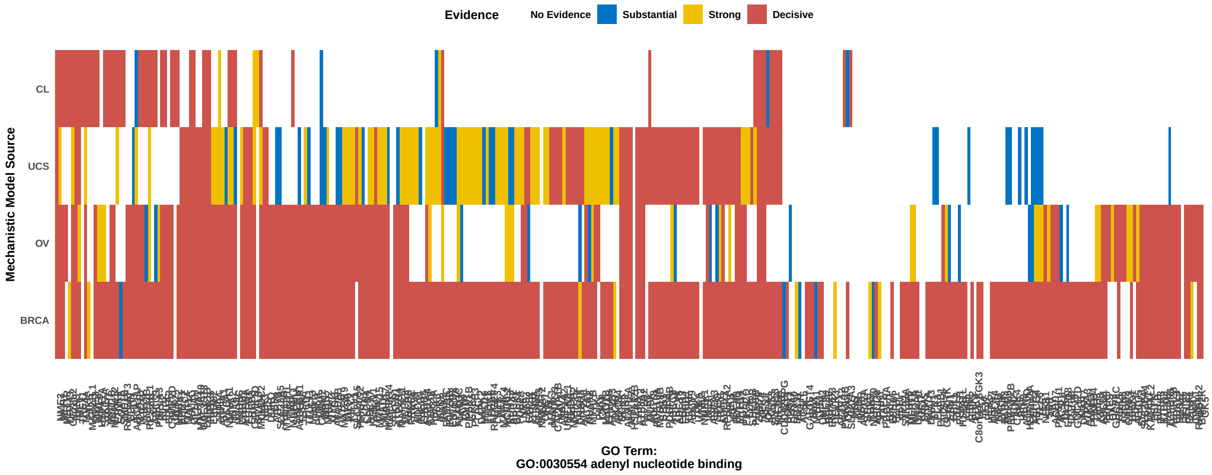


**Supplementary Figure S22: Heatmap summarizing levels of mechanistic evidence for the genes in GO adenyl neucleotide binding gene set.** Genes in the rows are ordered based on clusters resulting from the evidence statistics.

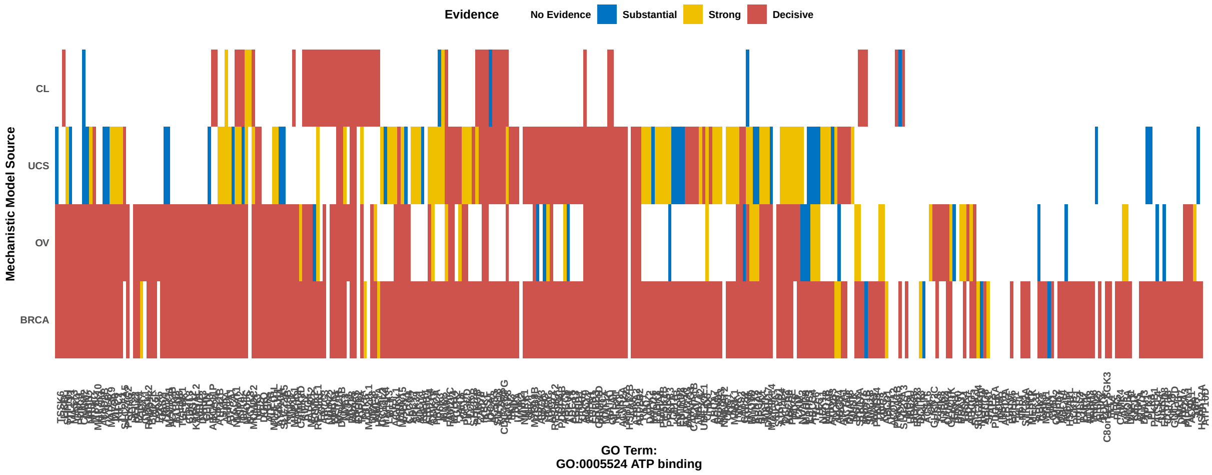


**Supplementary Figure S23: Heatmap summarizing levels of mechanistic evidence for the genes in GO ATP binding gene set.** Genes in the rows are ordered based on clusters resulting from the evidence statistics.

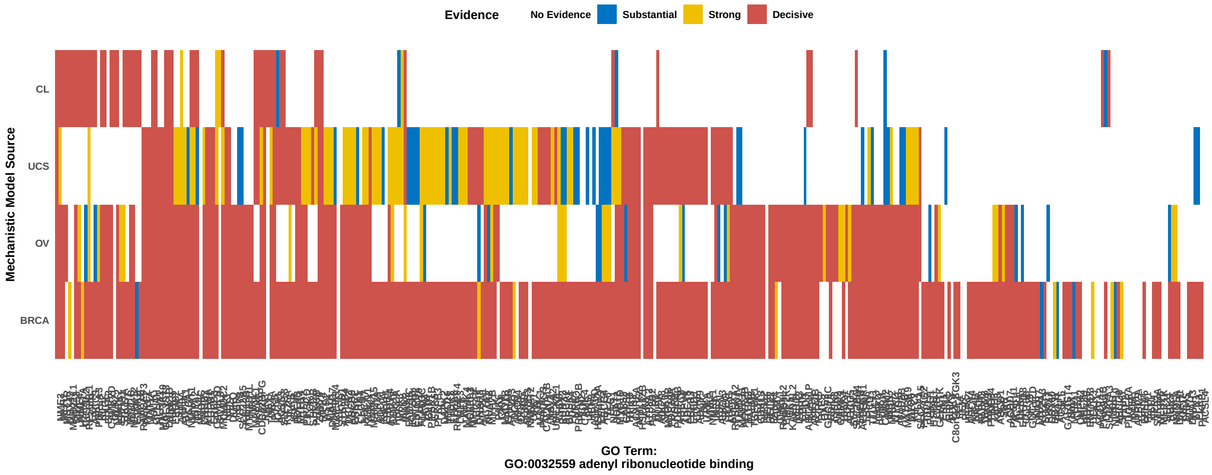


**Supplementary Figure S24: Heatmap summarizing levels of mechanistic evidence for the genes in GO adenyl riboneucleotide binding gene set.** Genes in the rows are ordered based on clusters resulting from the evidence statistics.

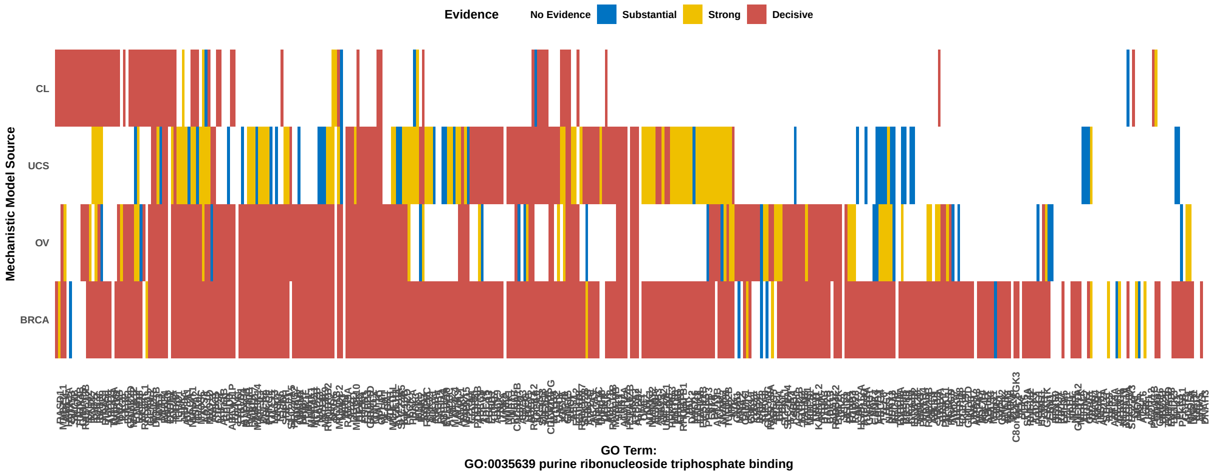


**Supplementary Figure S25: Heatmap summarizing levels of mechanistic evidence for the genes in GO purine riboneucleotide triphosphate gene set.** Genes in the rows are ordered based on clusters resulting from the evidence statistics.

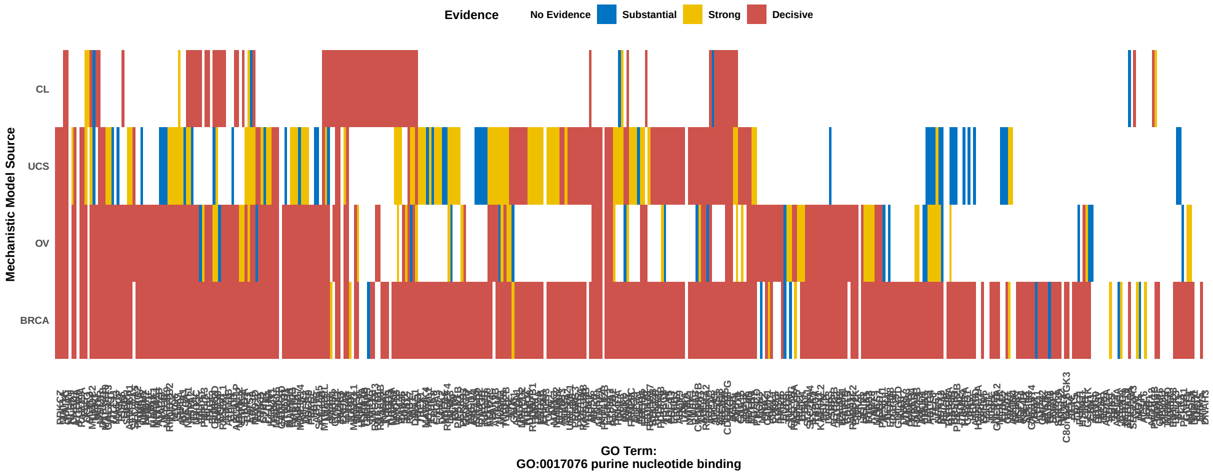


**Supplementary Figure S26: Heatmap summarizing levels of mechanistic evidence for the genes in GO purine neucleotide gene set.** Genes in the rows are ordered based on clusters resulting from the evidence statistics.

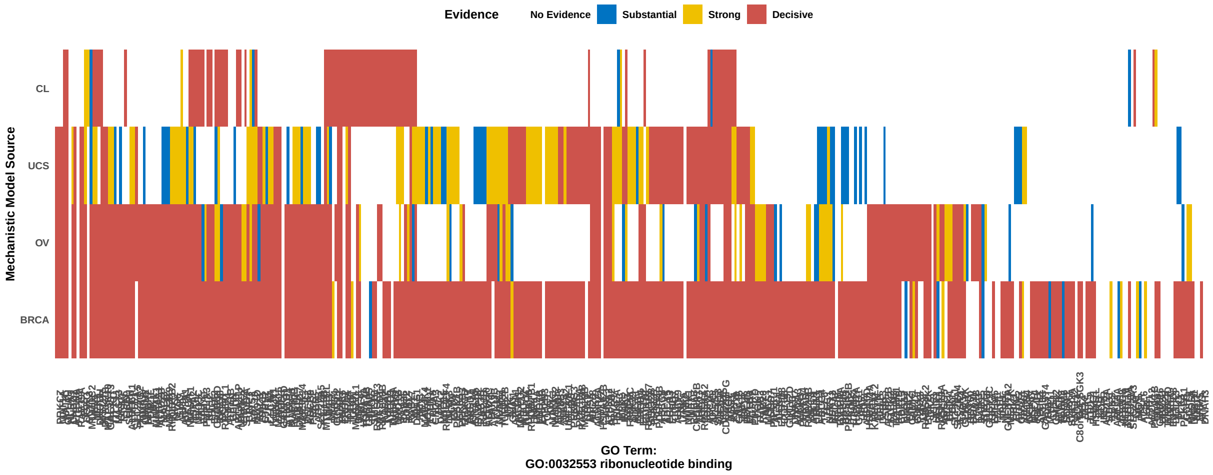


**Supplementary Figure S27: Heatmap summarizing levels of mechanistic evidence for the genes in GO riboneucleotide binding gene set.** Genes in the rows are ordered based on clusters resulting from the evidence statistics.

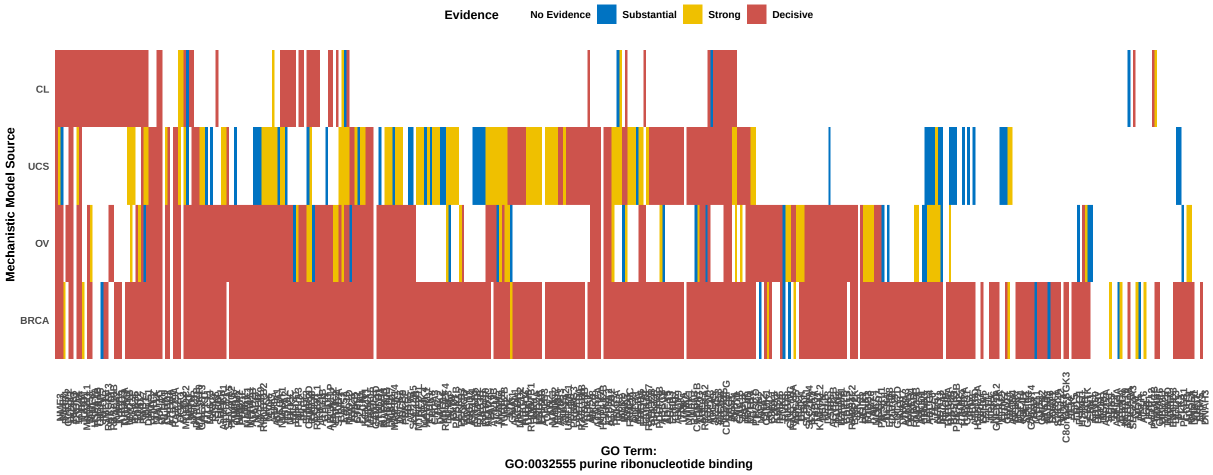


**Supplementary Figure S28: Heatmap summarizing levels of mechanistic evidence for the genes in GO purine riboneucleotide binding gene set.** Genes in the rows are ordered based on clusters resulting from the evidence statistics.

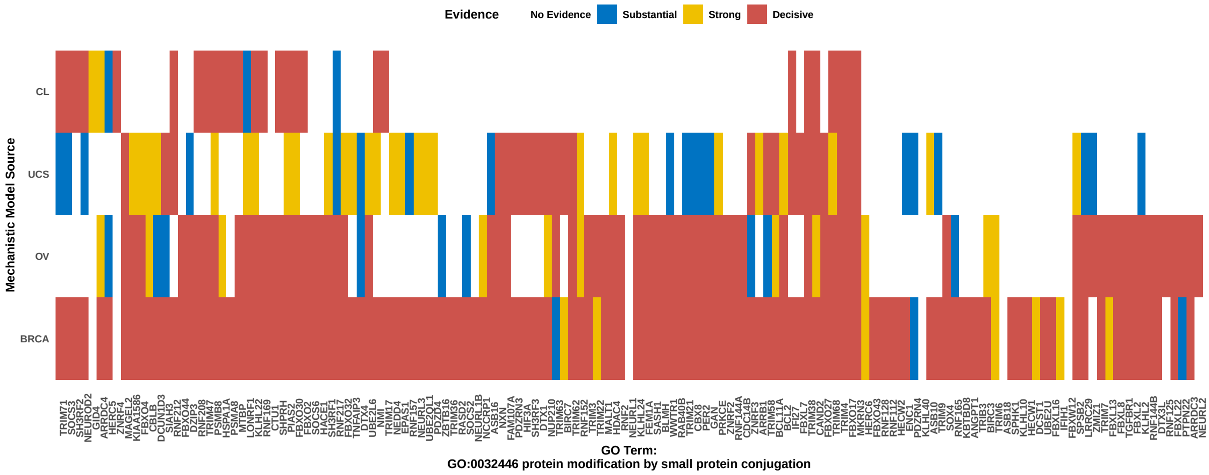


**Supplementary Figure S29: Heatmap summarizing levels of mechanistic evidence for the genes in GO protein modification by small protein conjugation gene set.** Genes in the rows are ordered based on clusters resulting from the evidence statistics.
